# Mathematical Analysis of a Zika Model with reservoirs and Human Movement

**DOI:** 10.1101/2022.03.02.22271760

**Authors:** Kifah Al-Maqrashi, Fatma Al-Musalhi, Ibrahim M. Elmojtaba, Nasser Al-Salti

## Abstract

A mathematical model for Zika virus is proposed describing the spread of the disease in three interacting populations, namely, human, vector (mosquitoes) and non-human primate (monkeys) inhabiting forests area. Human movement between rural and forest areas has been also considered. It is assumed that Zika virus spreads within non-human primate population, which in turn acts as a reservoir of infection, and then transmitted to the human population through infected mosquitoes. The proposed model incorporates vertical transmission and direct transmission in all populations. The proposed model has been first normalized. The normalized model has been then fully analyzed both qualitatively and quantitatively to investigate the role of the interaction between forest mosquitoes and primates on the ZIKV transmission dynamics. The mathematical analysis includes positivity and boundedness of solutions, derivation of the basic reproduction number *R*_0_ using the next generation matrix method, sensitivity analysis, existence and stability analysis of all equilibria and bifurcation analysis. Finally, numerical simulations have been carried out to illustrate the obtained theoretical results and to demonstrate the effect of some model parameters in the disease transmission dynamics. The results show that the interaction between forest mosquitoes and primates has a significant impact on the ZIKV transmission dynamics among human population through the fraction of susceptible moving to forest areas. Furthermore, the results highlight that the transmission probabilities are as important as the ratios of population size between vector population and human or primate populations in the disease transmission dynamics.

## Introduction

Recently, Zika virus infection has become a perilous danger for the human society [1]. It is a vector borne illness, spreads through the mosquito borne flavi-infection. This infection was first distinguished from rhesus monkeys in 1947 in the Zika forest of Uganda and from people in Nigeria in 1954, yet it did not spread in pestilence structure among the human populace until 2007 [1]. During 2013-2014, the largest epidemic of Zika virus occurred in Polynesia, France [2]. In 2018, the first principal assembled report of the biggest announced Zika outbreak in India [3]. Although, instances of Zika infection have fallen in the Americas, the Zika infection stays an active threat in certain areas of the world.

The infection is basically spread by the bites of Aedes species mosquitoes. Also, different types of transmission routes have been perceived (e.g., sexually and vertically). Generally, Zika infection causes a non-severe disease, i.e., it was characterized by causing mild symptoms like fever, headache, rash, arthralgia and conjunctivitis. However, a few territories recently influenced by the infection are giving troubling data on the all-around referenced possible relationship of neonatal mutations (microcephaly) and Guillian-Barre syndrome (GBS) with Zika Virus (ZIKV). As an outcome, the WHO pronounced a Public Health Emergency of International Concern on the first of February of 2016, featuring the significance of upgrade the actions to decrease the ZIKV infection, especially among pregnant ladies and ladies of childbearing age [4]. The advancement of Zika immunization will require proceeded with center and speculation. Until a Zika antibody is accessible, counteraction endeavors for pregnant ladies incorporate aversion of movement to territories with dynamic Zika transmission, evasion of mosquito bites for those living in or going to zones with Zika transmission, and insurance against sexual transmission [5].

Researchers from different fields are working on understanding the disease transmission dynamics. Mathematical models have been demonstrated as a significant tool to comprehend the disease infection transmission. However, there have been few studies on the role of animals as ZIKV hosts. There are some reports on ZIKV identification in non-human primates which raise the prospect that they could serve as reservoirs. The creation of a sylvatic Zika cycle in the forest of South America is highly likely, according to mathematical models of Zika virus transmission as discussed in [6]. Their model predicts that the inclusion of a quickly reproducing primate or other mammal, that is a competent host for ZIKV, greatly boosts the possibilities of establishing a sylvatic cycle. They suggest that the formation of a ZIKV sylvatic cycle can be supported by a network of as few as 6,000 primates and 10,000 mosquitoes. Furthermore, during the 2011 ZIKV amplification in Kedougou, field researchers revealed that the virus was present in all main land cover classes in the region, but was discovered substantially more frequently in the forest than in other land cover types. Two of the three monkey species found in Kedougou, African green monkeys and Patas monkeys, have previously been found to carry ZIKV. Following that, the virus was discovered in a large swath of tropical Africa using monkey serosurveys and virus isolation from monkeys and many species of sylvatic Aedes. In [7], scientists predict that when monkeys become infected with the sylvatic cycle of Yellow Fever (also a flavivirus) in the Americas, they show overt clinical symptoms and a viremia high enough to transmit virus to mosquito vectors. The virus remained limited to Africa and Asia’s tropical areas, infecting monkeys, arboreal mosquitoes, and humans on rare occasions. The majority of primates found to be ZIKV-positive in the wild or in sentinel studies are from the Old World. Humans are more closely linked to Old World primate species, particularly chimps and orangutans, according to phylogenetic study. Hence, the relative risk often increase in diseases that can be transmitted between closely related species. They also discovered that the ZIKV genome sequence in monkeys was identical to the ZIKV circulating in humans in South America, implying that primates sharing the same habitat as humans could serve as ZIKV hosts. In addition, scientists in [8] look into ZIKV infection in wild African monkeys to learn more about its emergence and transmission, and see if there was any signs of active or prior infection. Their findings imply that up to 16% of some nonhuman primate groups were exposed to ZIKV at some stage.

Researchers in [9] predict that studying ZIKV in Non-Human Primates (NHP) models can provide insight into viral dynamics as well as serve as a valuable tool for testing antiviral medicines and vaccines. Many of the important clinical findings in human Zika infection have been replicated in NHPs, including quick control of acute viremia, early penetration of the central nervous system, and sustained viral shedding and fetal disease in pregnant animals.

Moreover, in [10], a ZIKV mathematical model incorporating human movement between rural areas and nearby forests was presented to investigate the role of human movement in the spread of Zika virus infections in human and vector populations. The vector compartment, based on mosquito species distributions, have been split into rural areas and proximity forest areas. Authors discussed the consequences of an infected person with mild symptoms moving from rural to nearby forest regions in search of work or sustenance. The effect of a susceptible human mobility on the ZIKV transmission has been also considered. The obtained results showed that the human movement from rural areas to forests has a small effect in increasing the infected human and vector populations.

In this paper, we extend our previous work by proposing a mathematical model that includes three interacting populations, namely, human, mosquitoes (vectors) and monkeys (primates). The vector population has been divided into rural and forest mosquitoes. Direct and vertical transmissions routes are proposed in all populations. The proposed model aims to assess the effect of non-human primate (monkeys) inhabiting forests in spread of ZIKV to the nearby rural areas through the mobility of susceptible humans to forest areas and their interaction with contagious forest mosquitoes. This nearby movement can be happen for different reasons like work or food searching. The rest of this paper is organized as follows: model description is given in Section 2. Mathematical Analysis of the proposed model is discussed in Section 3. The Mathematical analysis includes normalization, positivity and boundedness of solution, basic reproduction number, sensitivity analysis of reproduction number, local and global stability of equilibrium states and bifurcation analysis. Numerical simulations of the proposed model are presented in Section 4, which includes examining the effect of some model parameters variations on the disease transmission dynamics. Finally, all findings are concluded in Section 5.

### Model Description

In this section, a model for Zika virus transmission between humans, vectors and primates (monkeys) is proposed. The interactions between these different populations are illustrated in Fig 1. The total human population *N*_*H*_ is assumed to remain constant and is classified into three compartments: susceptible *S*_*h*_, infected *I*_*h*_ and recovered *R*_*h*_ such that *N*_*H*_ = *S*_*h*_ + *I*_*h*_ + *R*_*h*_. Recovered humans are assumed to have a lifelong immunity [2].

**Fig 1.**
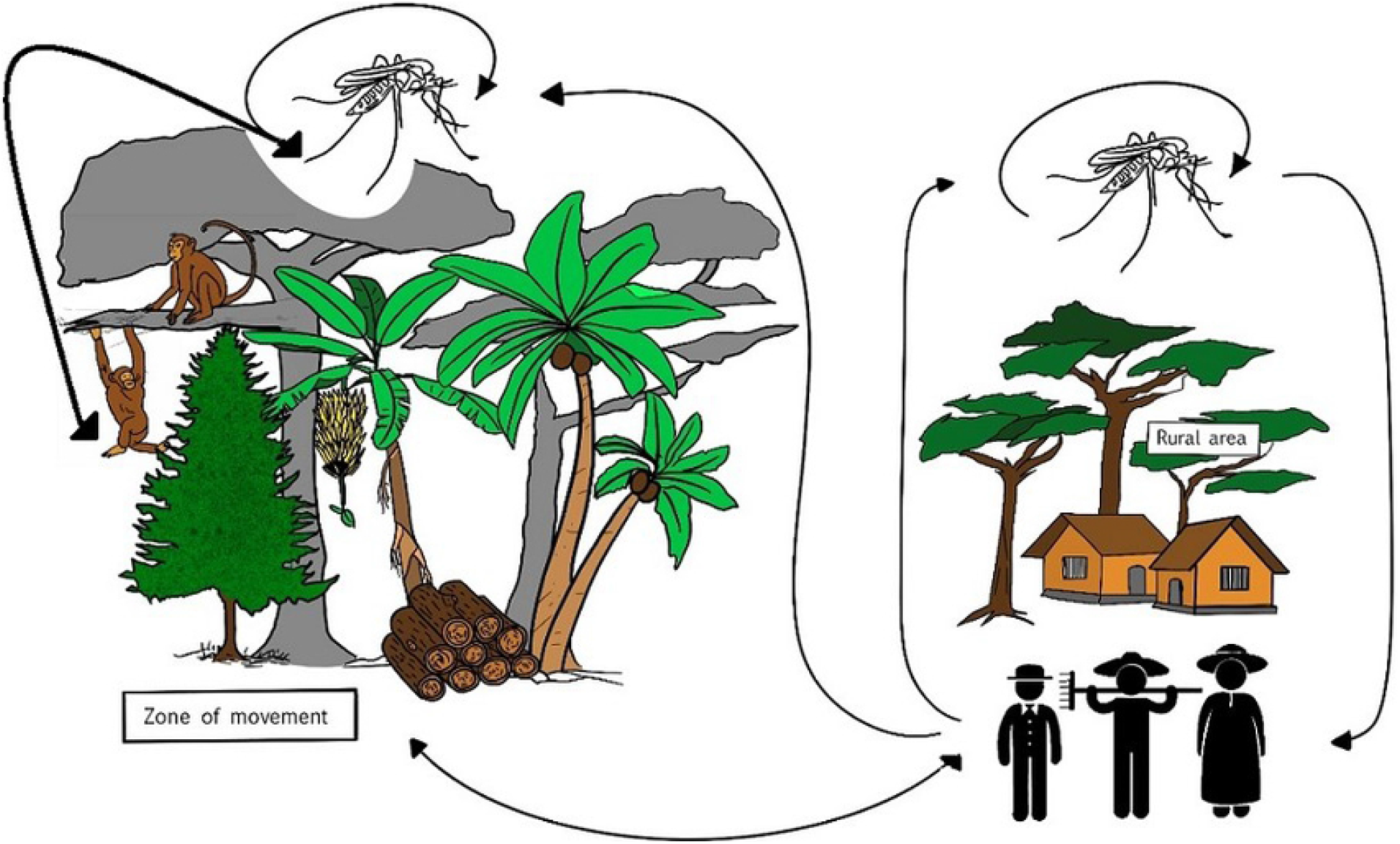
Illustrated figure for the interaction between human, vectors and non human primates populations.

The susceptible humans are assumed to gain the infection through bites of infectious female mosquitoes that live in rural areas *I*_*v*_ as well as by infectious female mosquitoes that live in forest alienated nearby rural areas *I*_*u*_ due to their movement to forest areas. So, we split the vector population according to its living area into: rural population (*S*_*v*_, *I*_*v*_) and nearby forest population (*S*_*u*_, *I*_*u*_) with constant total vector populations *N*_*V*_ and *N*_*U*_ such that *N*_*V*_ = *S*_*v*_ + *I*_*v*_ and *N*_*U*_ = *S*_*u*_ + *I*_*u*_, where *S*_*v*_ and *S*_*u*_ are the susceptible mosquitoes in rural and nearby forest areas, respectively. Recovery of infected mosquitoes from ZIKV infection is not considered due to its short life span [2]. They are assumed to remain infected for their entire remaining life. [6]. We assume that non-humans primates (reservoirs) are monkeys and are classified into three compartments *S*_*p*_, *Ip* and *R*_*p*_ similar to human population with constant total population *N*_*P*_ such that *N*_*P*_ = *S*_*p*_ + *I*_*p*_ + *R*_*p*_. They live only in forest areas and can get infected only by forest infectious mosquitoes. When monkeys become infected, they present overt clinical signs high enough to transmit virus to the mosquito vectors [7]. Monkeys primates recover at a fixed rate [6] and when challenged again with the virus do not get infected [11]. The description of the transmission of the ZIKV infection between these compartments are stated as follows:

i. Susceptible humans *S*_*h*_ can get infected with ZIKV via three main routes [12]: vector transmission by a female mosquito bite, direct transmission via sexual transmission or blood transfusion, or vertical transmission by being passed from mother to her newborn child. We assume that a fraction *ϵ*_1_ of newborns to infected are affected by Zika. We assume that the Zika-affected newborns enter the infected class. Evidence suggests that this fraction is about 2*/*3 [13]. A fraction *κ* of the susceptible human individuals progresses from rural areas to the nearby forest area for specific reasons such as work or food searching. So, they may get infection from infectious mosquitoes in forest area as well.
ii. Susceptible rural mosquitoes *S*_*v*_ can only be infected by infectious humans *I*_*h*_ and susceptible forest mosquitoes *S*_*u*_ can only be infected by infectious primates *I*_*p*_. Vertical transmission *ϵ*_2_, *ϵ*_3_ of the Zika virus in both vector populations are incorporated, respectively. Evidence suggests that Zika virus is transmitted vertically in the mosquito vector [14] and this is the main pathway it survives the colder months.
iii. Susceptible primates *S*_*p*_ are infected by forest infectious mosquitoes and infectious primates *I*_*p*_ can infect forest susceptible mosquitoes *S*_*u*_ as well. We assume symptomatic and asymptomatic primates equally infectious to mosquitoes and we lump the two classes in one, *I*_*p*_. A proportion *ϵ*_4_ of primates population can transmit ZIKV infection vertically. Infectious monkeys can also infect other susceptible partner sexually.

The above description is illustrated in Fig 2 and accordingly, the proposed model is given by the following set of equations:

**Fig 2.**
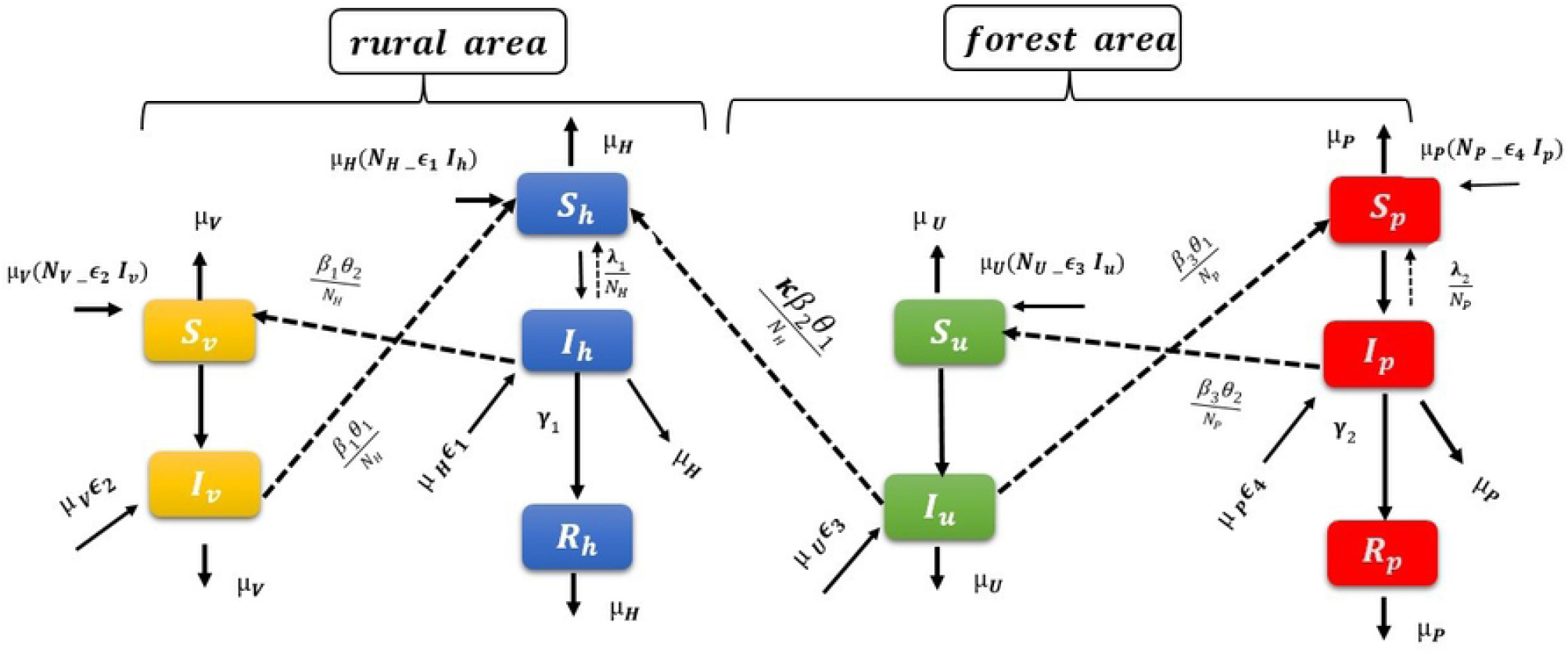
Progression diagram of the of the proposed ZIKV model.

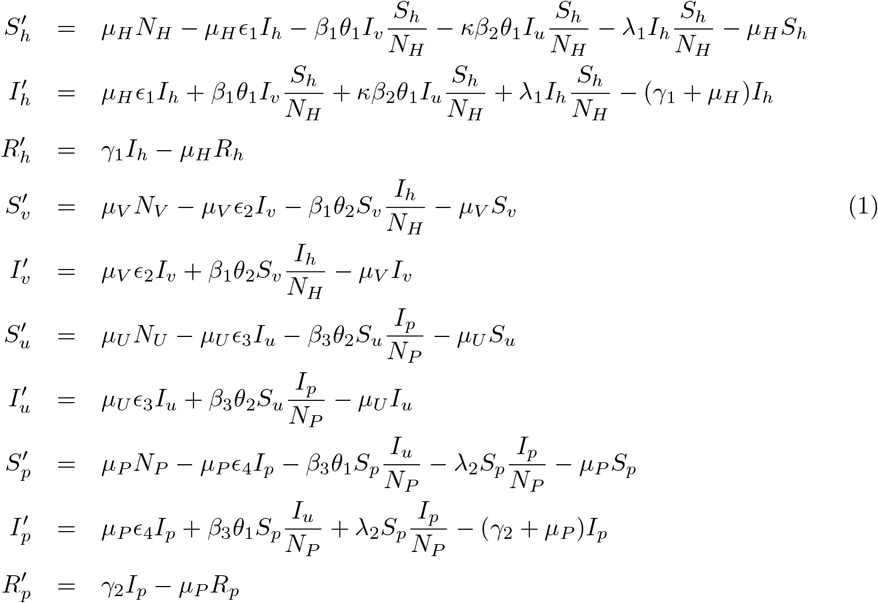

subjected to non-negative initial conditions

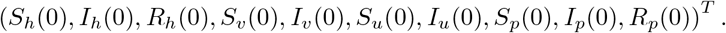

The parameter values of the model (1) is given in Table 1.

**Table 1.**
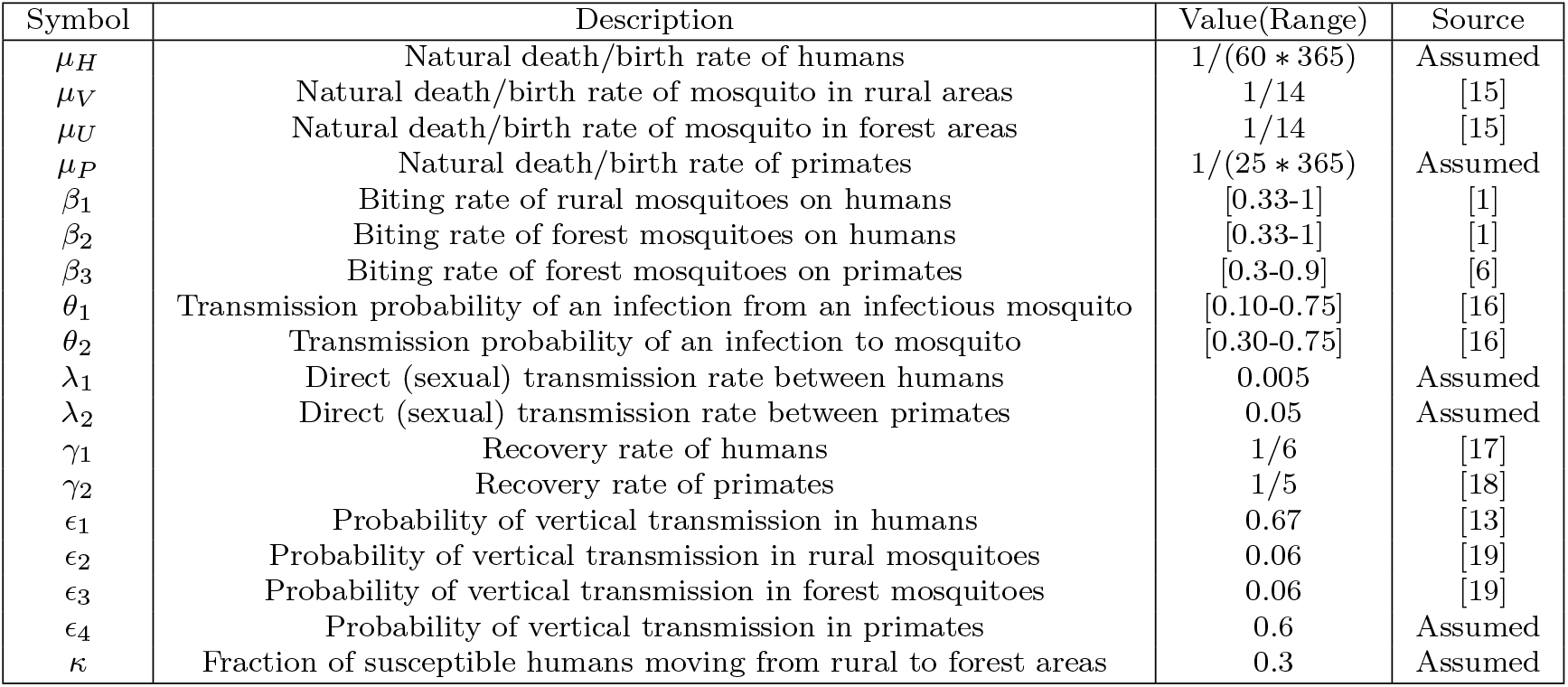
Description of the constant parameters used in model (1)and its values (per day).

| Symbol | Description | Value(Range) | Source |
| --- | --- | --- | --- |
| $\mu_H$ | Natural death/birth rate of humans | 1/(60 * 365) | Assumed |
| $\mu_V$ | Natural death/birth rate of mosquito in rural areas | 1/14 | [15] |
| $\mu_U$ | Natural death/birth rate of mosquito in forest areas | 1/14 | [15] |
| $\mu_P$ | Natural death/birth rate of primates | 1/(25 * 365) | Assumed |
| $\beta_1$ | Biting rate of rural mosquitoes on humans | [0.33-1] | [1] |
| $\beta_2$ | Biting rate of forest mosquitoes on humans | [0.33-1] | [1] |
| $\beta_3$ | Biting rate of forest mosquitoes on primates | [0.3-0.9] | [6] |
| $\theta_1$ | Transmission probability of an infection from an infectious mosquito | [0.10-0.75] | [16] |
| $\theta_2$ | Transmission probability of an infection to mosquito | [0.30-0.75] | [16] |
| $\lambda_1$ | Direct (sexual) transmission rate between humans | 0.005 | Assumed |
| $\lambda_2$ | Direct (sexual) transmission rate between primates | 0.05 | Assumed |
| $\gamma_1$ | Recovery rate of humans | 1/6 | [17] |
| $\gamma_2$ | Recovery rate of primates | 1/5 | [18] |
| $\epsilon_1$ | Probability of vertical transmission in humans | 0.67 | [13] |
| $\epsilon_2$ | Probability of vertical transmission in rural mosquitoes | 0.06 | [19] |
| $\epsilon_3$ | Probability of vertical transmission in forest mosquitoes | 0.06 | [19] |
| $\epsilon_4$ | Probability of vertical transmission in primates | 0.6 | Assumed |
| $\kappa$ | Fraction of susceptible humans moving from rural to forest areas | 0.3 | Assumed |

Model Mathematical Analysis In this section, we first normalize the proposed model (1). Then, we study positivity of solutions and invariant set, the basic reproduction number and its sensitivity analysis. Existence of equilibrium points, their local and global stability and bifurcation analysis are also considered.

### Model Normalization

To normalize the proposed model (1), let

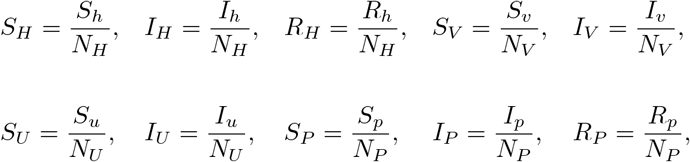

such that

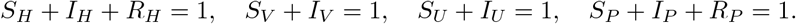

Thus, the normalized model is given by:

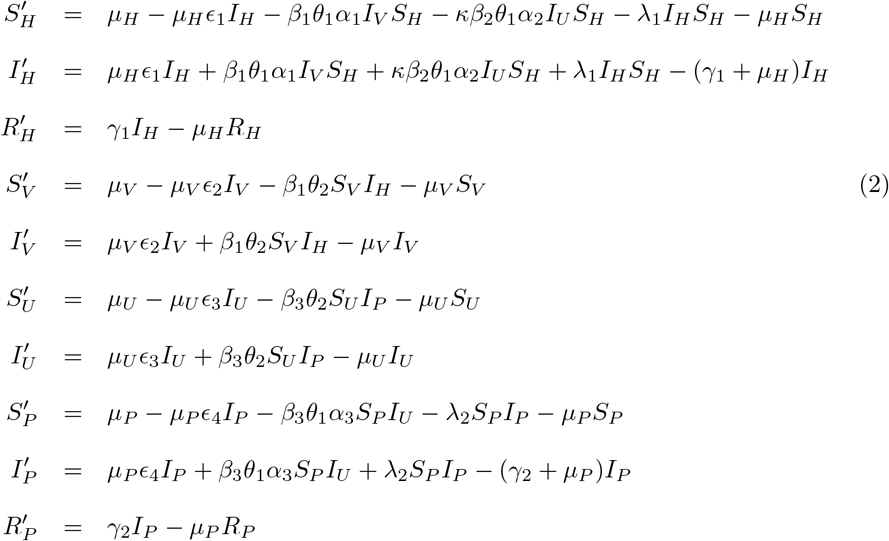

where 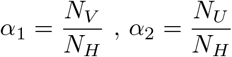 and 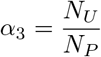 with non negative initial condition

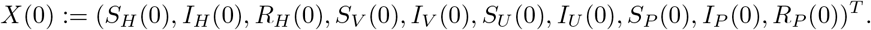

### Positivity of Solutions and Positively Invariant Set

The positivity and boundedness of the state variables are demonstrated in the following theorem :

#### Theorem 1

*The solution*

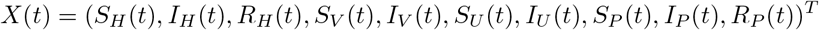

*of system (2) with non-negative initial condition X*(0) *remains positive for all time t* > 0 *in a positively invariant closed set*

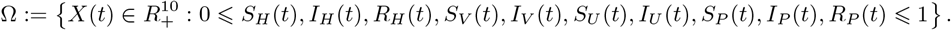

**proof**

Assume that the system (2) has non-negative initial condition *X*(0). Let *τ* = *sup*{*t* > 0; *X*(*t*) > 0}. Then, 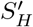 can be written as

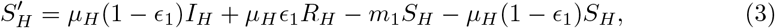

where, *m*_1_ = *β*_1_*θ*_1_*α*_1_*I*_*V*_ + *κβ*_2_*θ*_1_*α*_2_*I*_*U*_ + *λ*_1_*I*_*H*_ > 0. It follows that

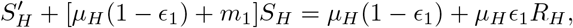

then integrating both sides over (0, *τ*), we get

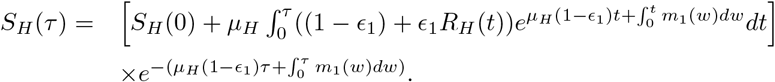

Clearly, *S*_*H*_(*τ*) is positive since *S*_*H*_(0) ≥ 0. Similar calculations can be done for 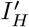 and 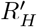, we get

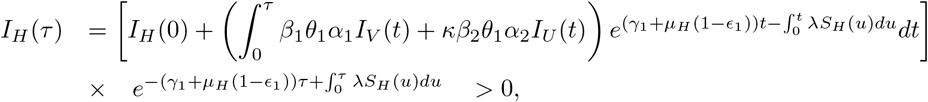

and

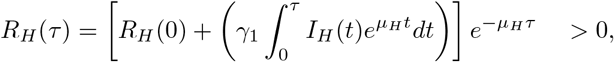

since initial conditions are non-negative. Similarly, one can show that the remaining components of *X*(*t*) are positive at *τ*. Using continuity of solution and *X*(0) ≥ 0, we conclude that *τ* can not be supremum and hence, solution will remain positive for all *t* > 0.

Now, assume that Ψ(*t*) = (Ψ_1_(*t*), Ψ_2_(*t*), Ψ_3_(*t*), Ψ_4_)^*T*^, where,

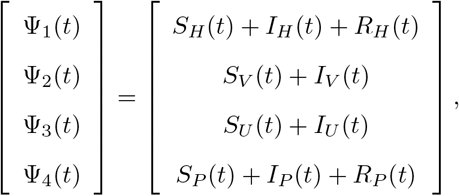

then, we have

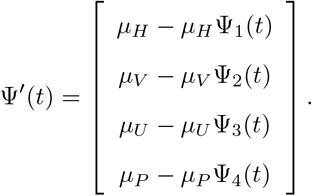

Now, solving for each component, we get :

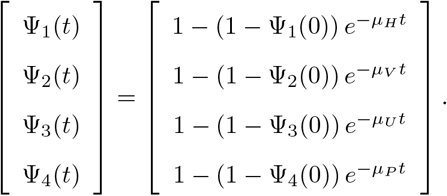

where Ψ_1_(0) = *S*_*H*_ (0) + *I*_*H*_ (0) + *R*_*H*_ (0), Ψ_2_(0) = *S*_*V*_ (0) + *I*_*V*_ (0), Ψ_3_(0) = *S*_*U*_ (0) + *I*_*U*_ (0), and Ψ_4_(0) = *S*_*P*_ (0) + *I*_*P*_ (0) + *R*_*P*_ (0). It is straight forward to say that

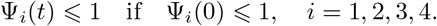

Thus, the set Ω is positively invariant set since 0 ⩽ Ψ(*t*) ⩽ 1. Moreover, if Ψ(0) > 1 then 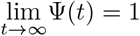 and therefore the set Ω is a globally attractive set.

### The Basic Reproduction Number

The model (2) has three different types of equilibria: disease free equilibrium (*DFE* : *Ƶ*^0^), axial equilibrium (*AE* : *Ƶ*^(1)^), describes an endemic in populations living in rural area only and endemic equilibrium (*EE* : *Ƶ*^(2)^), describes an endemic in both forest and rural areas. The endemic and axial equilibria will be discussed later.

The disease free equilibrium always exists and is given by

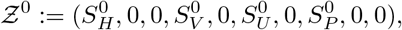

where, 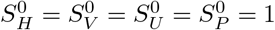. The threshold quantity ℛ_0_ is defined as the average number of secondary infections generated by one case in a completely susceptible population. It is calculated by well known Next Generation Method [20]. The Jacobian of the transmission matrix is:

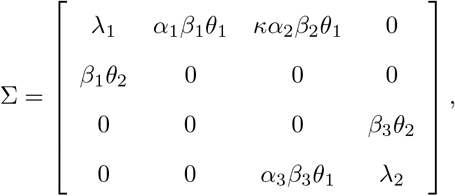

and Jacobian of the transition matrix is

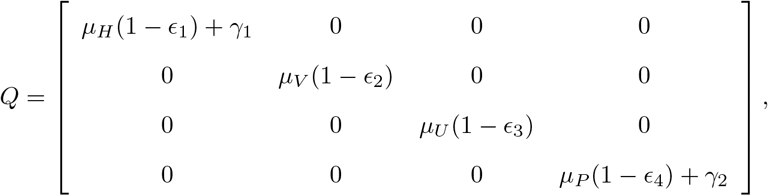

then, the next generation matrix Σ*Q*^*−*1^ is

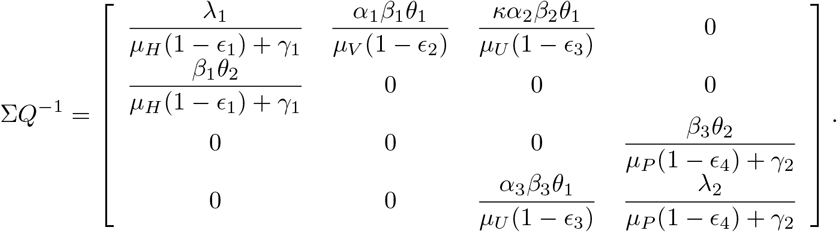

The basic reproduction number ℛ_0_ is the spectral radius of Σ*Q*^*−*1^ which can be expressed as:

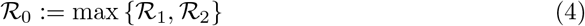

where

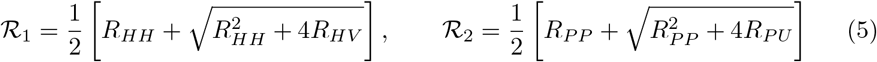

with 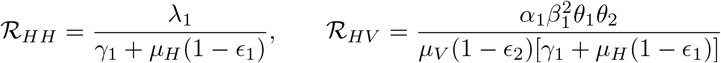, and 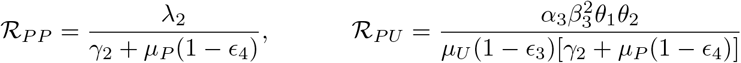.

It is clear that ℛ_*HH*_ and ℛ_*PP*_ represent the transmission due to interaction between human population and between primates population, respectively. ℛ_*HV*_ and ℛ_*PU*_ represent the transmission due to interaction between human and vector in rural areas and interaction between primates and vector in forest areas, respectively. Moreover, the disease threshold occurs at ℛ_0_ = 1 which implies either ℛ_1_ = 1 whenever ℛ_*HH*_ + ℛ_*HV*_ = 1 or ℛ_2_ = 1 whenever ℛ_*PP*_ + ℛ_*PU*_ = 1. Also, it can be easily proven that the disease infection will die out at ℛ_0_ *<* 1 which implies that ℛ_*HH*_ + ℛ_*HV*_ *<* 1 and ℛ_*PP*_ + ℛ_*PU*_ *<* 1. This will happen by reducing all routes of transmission, ℛ_*HH*_, ℛ_*HV*_ ℛ_*PP*_, and ℛ_*PU*_. However, the disease infection persists in all populations when ℛ_0_ > 1, whenever ℛ_*HH*_ + ℛ_*HV*_ > 1 and ℛ_*PP*_ + ℛ_*PU*_ > 1.

### Sensitivity Analysis of the Basic Reproduction Number

In this section, sensitivity analysis of the basic reproduction number ℛ_0_ is discussed. It allows us to determine the parameters which have significant impact on threshold ratio ℛ_0_ of model (2),i.e, a small change in a highly sensitive parameter leads to a high quantitative variation in ℛ_0_ and may produce qualitatively different results. Such parameters must deserve the attention of management and control strategies. Here, the normalized forward sensitivity index method (elasticity index) [21] is used and it is defined as the ratio of the relative change of ℛ_0_ with respect to relative variation in a parameter *l* as follows

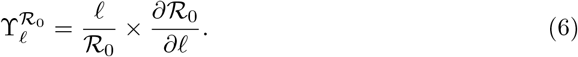

Since ℛ_0_:= max {ℛ_1_, ℛ_2_}, the sensitivity analysis of ℛ_0_ with respect to each of its parameter will be evaluated via the sensitivities of each of ℛ_1_ and ℛ_2_ such that

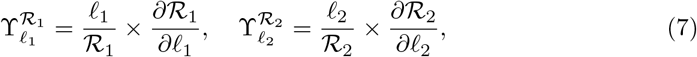

where *l*_1_, *l*_2_ denotes the parameter related to ℛ_1_ and ℛ_2_, respectively. Using the explicit expression of the basic reproduction number ℛ_0_ (5) with the base line values of parameters listed in Table 1, the estimated values of the sensitivities indices at ℛ_0_ > 1 are listed in Table 2.

**Table 2.** Sensitivity indices of ℛ_1_ and ℛ_2_.

| $\mathcal{R}_0 = \mathcal{R}_1 > \mathcal{R}_2 > 1$ | | | $\mathcal{R}_0 = \mathcal{R}_2 > \mathcal{R}_1 > 1$ | | |
| --- | --- | --- | --- | --- | --- |
| Parameter( $\ell_1$) | Sign | $\Upsilon_{\ell_1}^{\mathcal{R}_{01}}$ | Parameter( $\ell_2$) | Sign | $\Upsilon_{\ell_2}^{\mathcal{R}_{02}}$ |
| $\beta_1$ | + | 0.9889 | $\beta_3$ | + | 0.8775 |
| $\lambda_1$ | + | 0.0111 | $\lambda_2$ | + | 0.1224 |
| $\gamma_1$ | - | 0.5055 | $\gamma_2$ | - | 0.5611 |
| $\alpha_1$ | + | 0.4944 | $\alpha_3$ | + | 0.4387 |
| $\mu_H$ | - | 0.4513e-4 | $\mu_P$ | - | 0.0001234 |
| $\mu_V$ | - | 0.4944 | $\mu_U$ | - | 0.4387 |
| $\epsilon_1$ | + | 0.9164e-4 | $\epsilon_4$ | + | 0.0001851 |
| $\epsilon_2$ | + | 0.0315 | $\epsilon_3$ | + | 0.0280 |
| $\theta_1$ | + | 0.4343 | $\theta_1$ | + | 0.4387 |
| $\theta_2$ | + | 0.4343 | $\theta_2$ | + | 0.4387 |

The sensitivity indices are interpreted with regard to their magnitudes and signs. The basic reproduction number ℛ_0_ increases (decreases) with the increase (decrease) in those parameters that have positive sensitivity indices. Conversely, it increases (decreases) with the decrease (increase) in those parameters that have negative sensitivity indices. Moreover, the magnitude of sensitivity indices represents the relative importance of those parameters that drive the transmission mechanism of the disease. Clearly, when ℛ_0_ = ℛ_1_ the most efficacious parameters is the biting rate of rural mosquitoes on humans *β*_1_, which has a strong positive impact on the value of ℛ_0_. While the most efficacious parameter when ℛ_0_ = ℛ_2_ is the biting rate of the forest mosquitoes on primates *β*_3_. Furthermore, the results highlight that the transmission probabilities *θ*_1_, *θ*_2_ are as important as the ratios of population size *α*_1_, *α*_3_, in the disease transmission dynamics. Obviously, increasing the transmission probabilities will lead to an increase in ℛ_0_. There are many factors affecting the transmission probabilities such as the ZIKV dose in the blood-meal as shown in [22], where authors have demonstrated that increasing ZIKV dose in the blood-meal significantly increases the probability of mosquitoes becoming infected, and consequently disseminating virus and becoming infectious. On the other hand, the recovery rate of humans *γ*_1_ and the recovery rate of primates *γ*_2_ have the most negative sensitive indices. Also, the short life span of mosquitoes will reduce the spread of the disease and hence decrease in ℛ_0_. Thus, for controlling Zika virus, it is suggested to control the mosquito population and prevent their breeding near houses in rural area.

### Local Stability of the DFE

Here, we discuss the local stability of the disease free equilibrium *Ƶ*^0^ by finding the eigenvalues of the linearized system. The result is given in the following theorem:

#### Theorem 2

*The DFE, given by Ƶ* ^0^, *of the model (2) is locally asymptotically stable if* ℛ_0_ *<* 1. *Otherwise, it is unstable*.

**proof**

First, let us introduce the following:

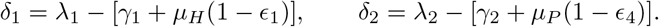

Then, the linearized matrix of the system (2) at the disease free equilibrium *Ƶ*^0^ is

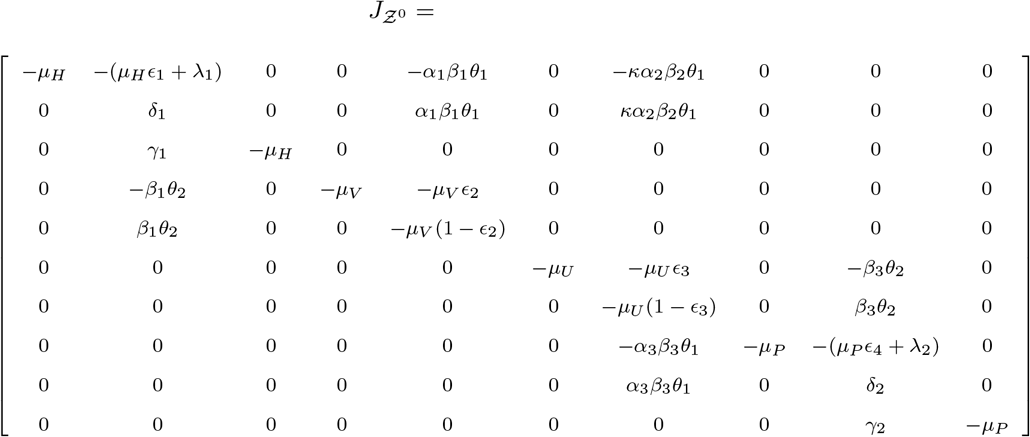

Clearly, the above system has six negative eigenvalues which are:

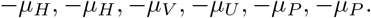

The remaining eigenvalues satisfy the characteristic equation *P* (*λ*) = 0, which is given by

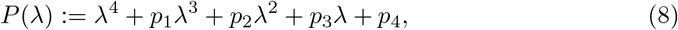

where

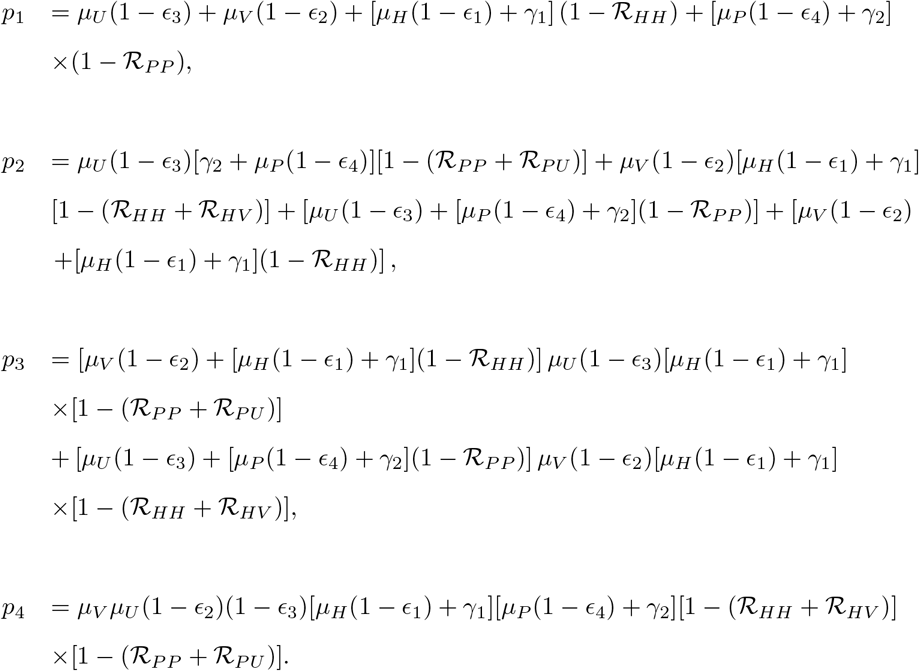

Obviously, *p*_4_ > 0 when ℛ_*HH*_ + ℛ_*HV*_ *<* 1 and ℛ_*PP*_ + ℛ_*PU*_ *<* 1 which implies that *p*_1_ > 0, *p*_2_ > 0 and *p*_3_ > 0. Hence, the necessary conditions of stability using Routh’s stability criterion [23] are satisfied. It is left to show that the sufficient conditions, namely, *p*_1_*p*_2_ − *p*_3_ > 0 and 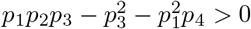 are satisfied. This can be done by computing

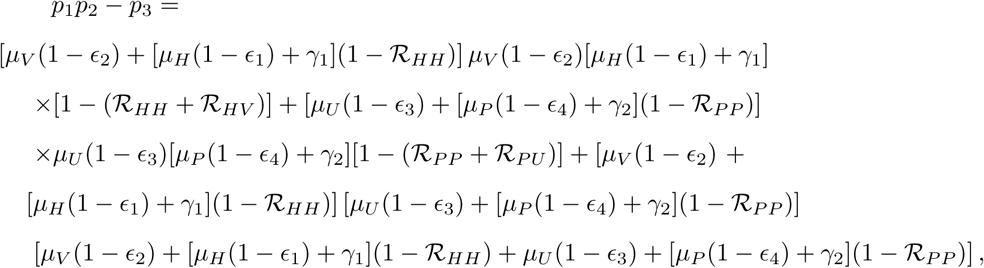

and

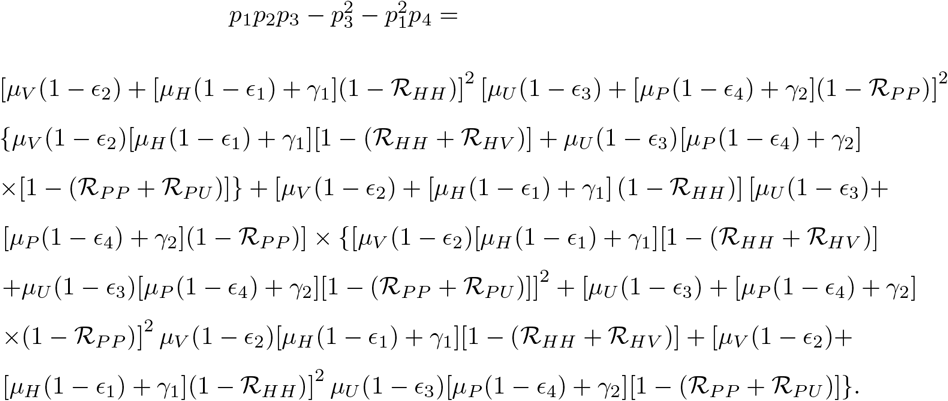

Clearly, *p*_1_*p*_2_ − *p*_3_ > 0 and 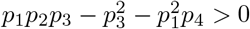 if and only if

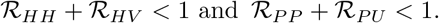

Hence, the eigenvalues of characteristic equation *p*(*λ*) = 0 have negative real parts whenever *R*_0_ *<* 1, which implies that the DFE (*Ƶ*^0^) is locally asymptotically stable.

### Global Stability of the DFE

The global stability of the DFE (*Ƶ*^0^) will ensure that the disease is eliminated under all initial conditions. In this regard, we state the following theorem:

#### Theorem 3

*The disease free equilibrium, given by Ƶ*^0^ *of the model (2) is globally asymptotically stable if R*_0_ *<* 1.

**proof**

The proof is based on using Castillo-Chavez theorem [24]. Let *X*(*t*) and *Y* (*t*) compartments describe the uninfected and infected classes of system (2), respectively:

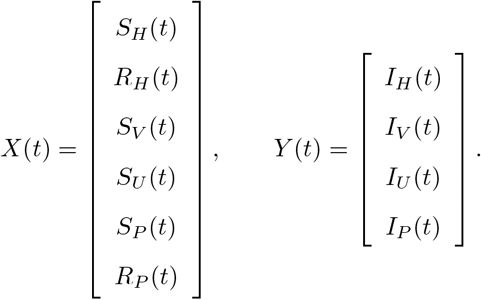

Therefore, system (2) can be written as :

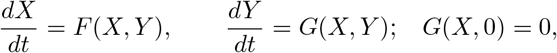

where *F* and *G* are the corresponding right hand sides in system (2).

According to Castillo-Chavez theorem, in order to guarantee the global asymptotic stability of the DFE (*Ƶ*^0^), the following two conditions (*H*1) and (*H*2) must be satisfied:

- (*H*1) *X*^0^= (1, 0, 1, 1, 1, 0)^*T*^ is globally asymptotically stable for 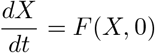.
- (*H*2) *Ĝ* ⩾ 0, where *Ĝ*(*X, Y*) = *AY* − *G*(*X, Y*) and *A* = *D*_*Y*_ *G*(*X*^0^, 0) is an Metzler matrix ∀(*X, Y*) ∈ Ω.

First to check (*H*1), we have

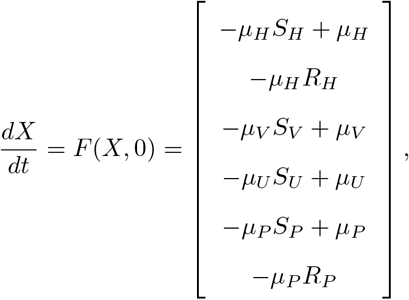

then, the behavior of each compartment can be obtained by solving the above system of ODEs and hence, we get

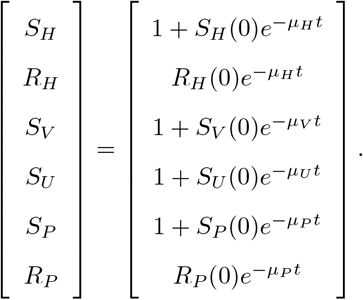

Clearly, 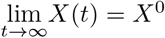, i.e, the first condition is satisfied. Next, we check the second condition (*H*2) by finding *A* = *D*_*Y*_ *G*(*X*^0^, 0) :

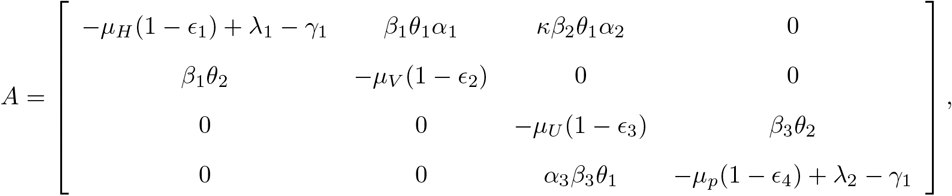

then, we compute *Ĝ*(*X, Y*) = *AY* − *G*(*X, Y*):

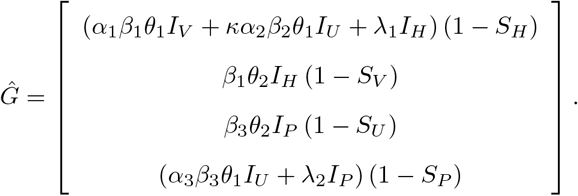

Clearly, *Ĝ* ⩾ 0, ∀(*X, Y*) ∈ Ω since 0 ⩽ (*S*_*H*_, *S*_*V*_, *S*_*U*_, *S*_*P*_) ⩽ 1. Thus, (*H*2) is satisfied.

Hence, *Ƶ*^0^ is globally asymptotically stable, provided that ℛ_0_ *<* 1.

### Existence of Endemic Equilibria

In this section, result related to the existence of axial and endemic equilibria is stated in the following theorem:

#### Theorem 4

*The model (2) poses two endemic equilibrium points* :

i. *a unique Axial equilibrium (AE) given by Ƶ* ^(1)^, *exists if* ℛ_1_ > 1,
ii. *a full Endemic equilibrium(EE) given by Ƶ* ^(2)^, *exists if* ℛ_2_ > 1.

**proof**

Let 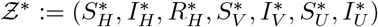 be the expected endemic equilibrium of the model (2). By solving 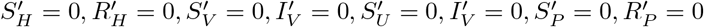, we get

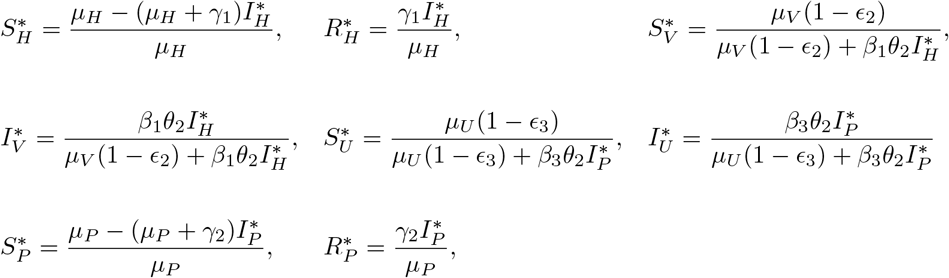

then by solving 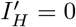 and using the expression of 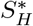, we get:

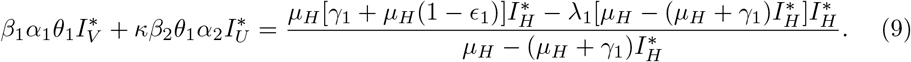

Substitution of 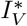 and 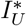 into equation (9) leads to the following equation in terms of 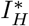 and 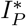:

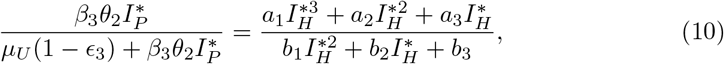

where

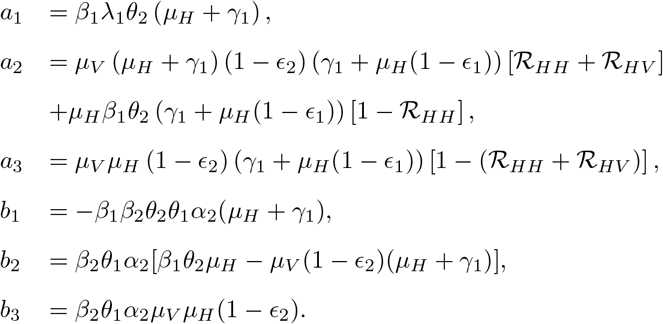

Solving 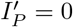 and using expressions of 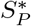 and 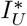 gives the following cubic equation:

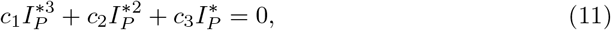

where

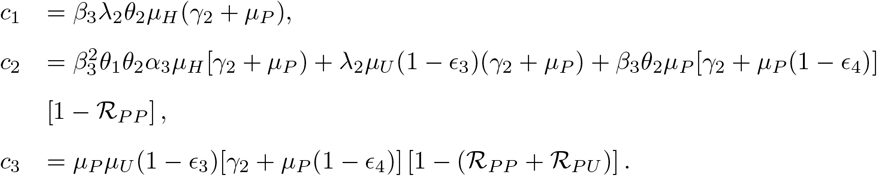

Now, from equation (11), we have the following two cases:

i. if 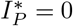, then equation (10) becomes a cubic equation in terms of 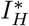:

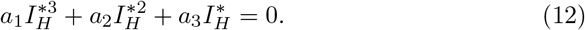

From the above equation, we have either 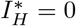 which gives the DFE or we get the quadratic equation:

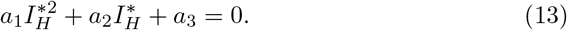

Clearly, *a*_1_ > 0 and *a*_3_ *<* 0 if and only if ℛ_*HH*_ + ℛ_*HV*_ > 1, which implies ℛ_1_ > 1. Thus, according to the Descartes’s Sign Rule [25], equation (13) has a unique positive root. Moreover, since 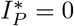, we have

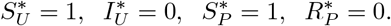

Hence, we have a unique AE, which is given by

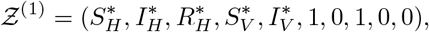

where,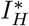 satisfies equation (13).
ii. If *I*_*P*_ ≠ 0, then solving equation (11) yields

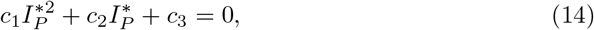

which has a unique positive root 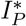 if ℛ_2_ > 1, since *c*_1_ > 0 and *c*_3_ *<* 0 if and only if ℛ_*PP*_ + ℛ_*PU*_ > 1.

Now, using the above result that 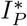 is the unique positive root of (14), equation (10) can be written terms of 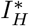 only:

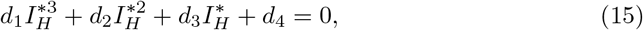

such that

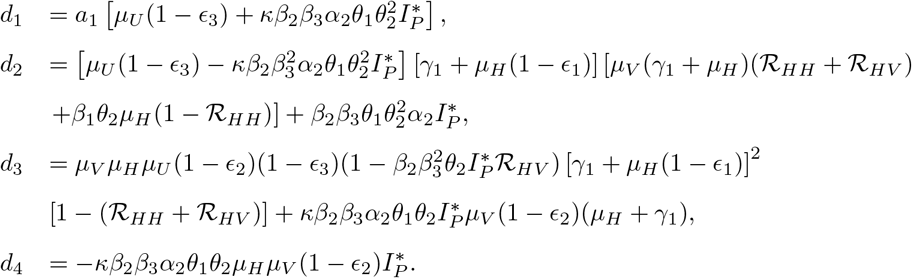

Hence, applying Descartes’s Sign Rule, provided that 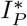 is positive if ℛ_2_ > 1, *d*_1_ > 0 and *d*_4_ *<* 0, we have the following possibilities for the existence of positive roots of equation (15):

I. a unique positive root, if *d*_2_ > 0 and *d*_3_ *<* 0 or both *d*_2_ and *d*_3_ have the same sign.
II. three positive roots, if *d*_2_ *<* 0 and *d*_3_ > 0.

Thus, EE exits if ℛ_2_ > 1 and can be written as

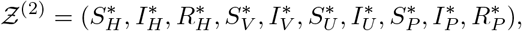

where, 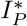 and 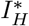 can be computed using equations (14) and (15), respectively.

### Bifurcation and Local Stability Analysis

Emergence or disappearance of new stable points causing a changes in stability of a system as a parameter changes is defined as bifurcation. In this section, we discuss the bifurcation of system (2) numerically and theoretically. All figures in this section is sketched using Matcont program [26].

Here, we take the transmission probability from an infectious mosquito, *θ*_1_, as a bifurcation parameter for which there are two possible bifurcations values,

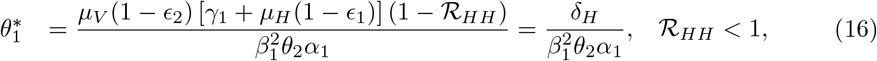

and

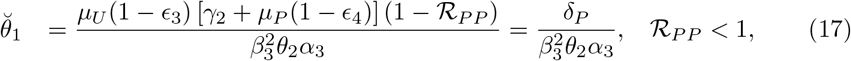

corresponding to ℛ_1_ = 1 and ℛ_2_ = 1, respectively, where *δ*_*H*_ = *µ*_*V*_ (1 − *ϵ*_2_)[*γ*_1_ + *µ*_*H*_ (1 − *ϵ*_1_)](1 − ℛ_*HH*_) and *δ*_*P*_ = *µ*_*U*_ (1 − *ϵ*_3_)[*γ*_2_ + *µ*_*P*_ (1 − *ϵ*_4_)](1 − ℛ_*PP*_).

Clearly, *θ*_2_ is the only common parameter between 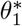 and 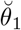. So, fixing the values of other parameters, 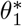 and 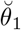 will attain their maximum values of one when

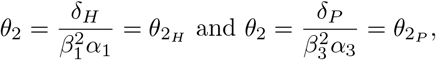

respectively, and their minimum values are obtained when *θ*_2_ = 1. Hence, the feasibility of the bifurcation values 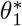 and 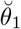 depends on the value of *θ*_2_. In particular, if the value of *θ*_2_ is taking to be less than 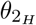 and 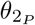, then both bifurcation values will be greater than one and thus no bifurcation will take place. The bifurcation of the system (2) will now be examined numerically for the following two cases:

a. ℛ_0_ = ℛ_1_ > ℛ_2_ (b) ℛ_0_ = ℛ_2_ > ℛ_1_ with *θ*_1_ as a bifurcation parameter, taking different values of *θ*_2_ and fixing the values of all other parameters. The bifurcation diagrams are illustrated in Figures 3 - 5.

In Figures 3 and 4, we assume that *β*_1_ = 0.35, *β*_2_ = 0.32, *β*_3_ = 0.3, *λ*_1_ = 0.1, and *λ*_2_ = 0.0025 such that *R*_0_ = ℛ_1_ > ℛ_2_. For these choices, we have 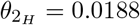 and 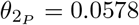. Choosing 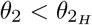, ℛ_0_ will be less than unity for all values of *θ*_1_ and hence the DFE is stable and no bifurcation will take place. Here, both 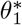 and 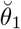 are not feasible. For example, when *θ*_2_ = 0.01, the critical values are 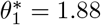 and 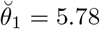. The bifurcation diagram for the case 0.0188 *< θ*_2_ *<* 0.0578 is illustrated in Fig 3. Here, the transmission probability of infection to mosquitoes is taken to be *θ*_2_ = 0.05 and the corresponding critical values are 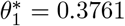 and 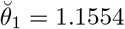, which is not feasible. Clearly, the system undergoes forward bifurcation when *θ*_1_ passes through the critical value 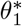 as shown in the Figure. Furthermore, when *θ*_2_ is taken to be 0.1, which is greater than both 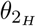 and 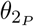, the critical values are 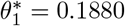and 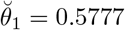. In this case, the system has double forward bifurcation when *θ*_1_ passes through these two critical values as shown in Fig 4. The system undergoes forward bifurcation at the DFE as *θ*_1_ passes through 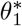 and at the AE as *θ*_1_ passes through 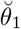.

**Fig 3.**
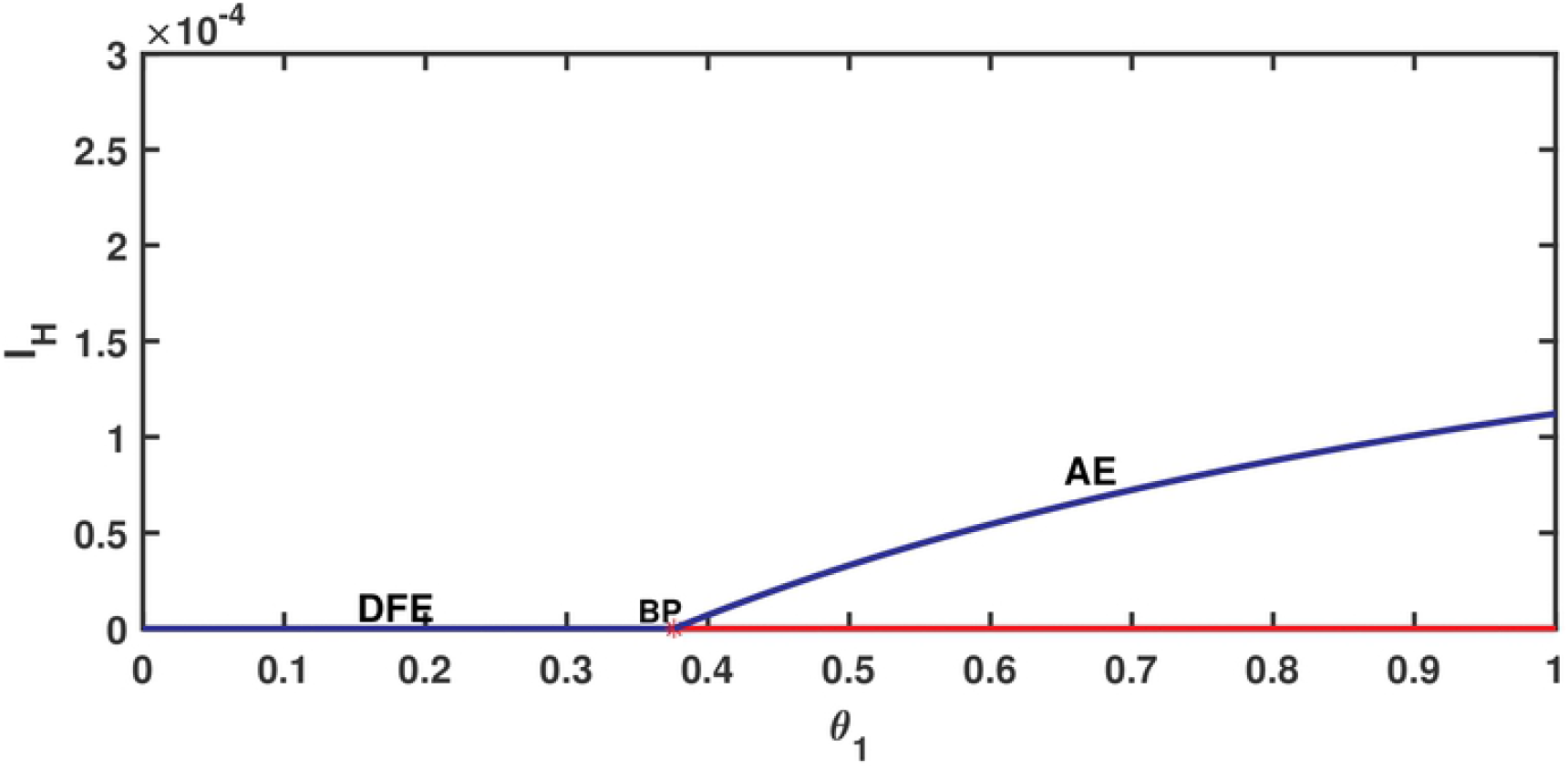
Forward bifurcation at the DFE with 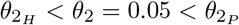 for the case ℛ_0_ = ℛ_1_ > ℛ_2_.

**Fig 4.**
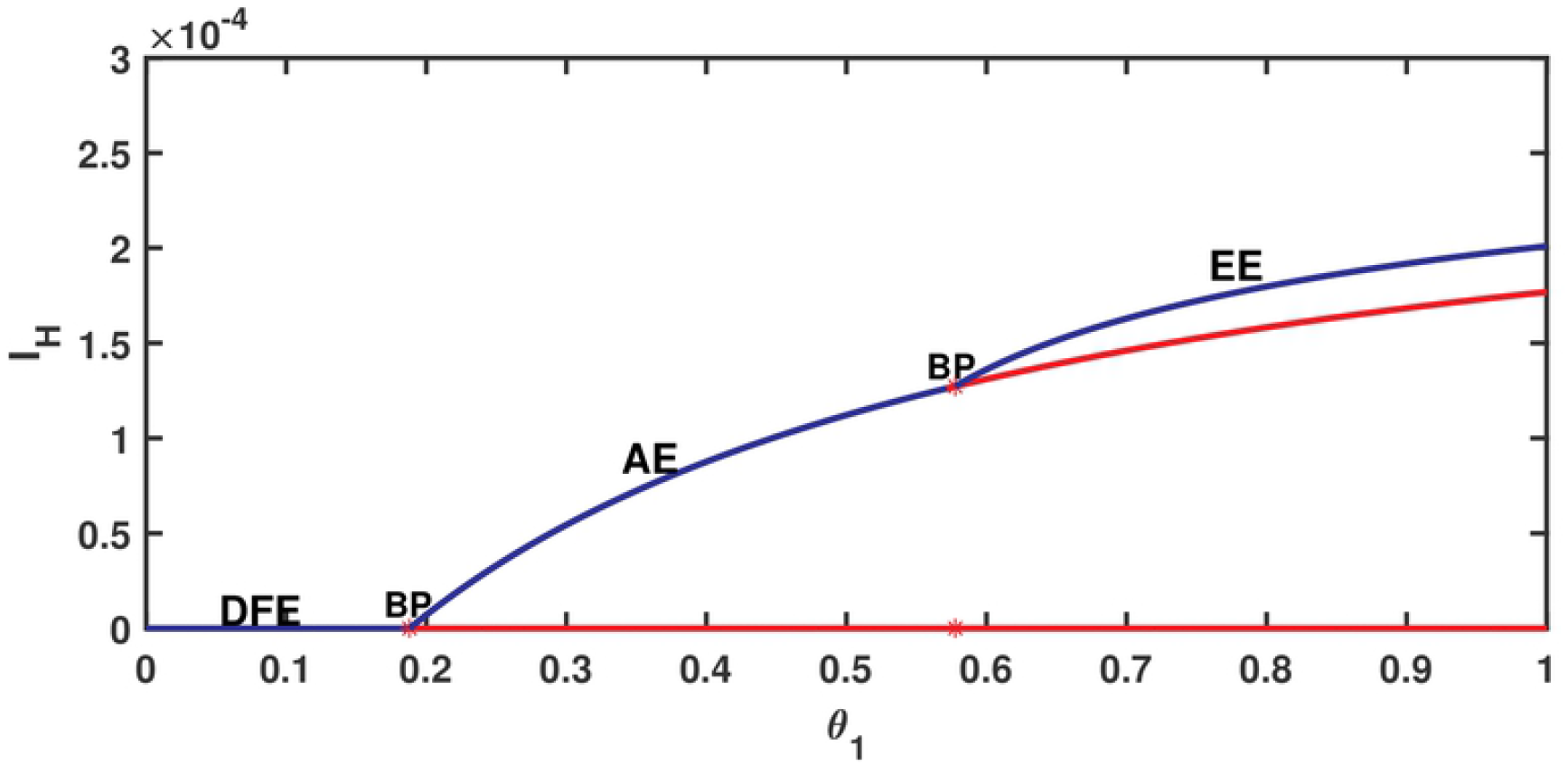
Double forward bifurcation at the DFE and AE, respectively, with 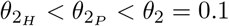 for the case ℛ_0_ = ℛ_1_ > ℛ_2_.

For the case *R*_0_ = ℛ_2_ > ℛ_1_, we choose *β*_1_ = 0.35, *β*_2_ = 0.32, *β*_3_ = 0.3, *λ*_1_ = 0.0025, and *λ*_2_ = 0.1, the corresponding values of 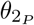 and 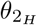 are 0.0293 and 0.0450, respectively. Similar to the previous case, the feasibility of the critical values of *θ*_1_ depends on the value of the *θ*_2_ and its relation to 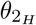 and 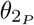. For illustration, we choose *θ*_2_ = 0.07 and the bifurcation diagram for this case is illustrated in Fig 5. The figure shows that the system has forward bifurcation at the DFE when *θ*_1_ passes through 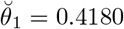 and that the EE once exists, it remains stable. Moreover, unstable AE exists when the system passes through the second critical value 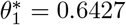 and hence there is no bifurcation at 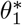.

**Fig 5.**
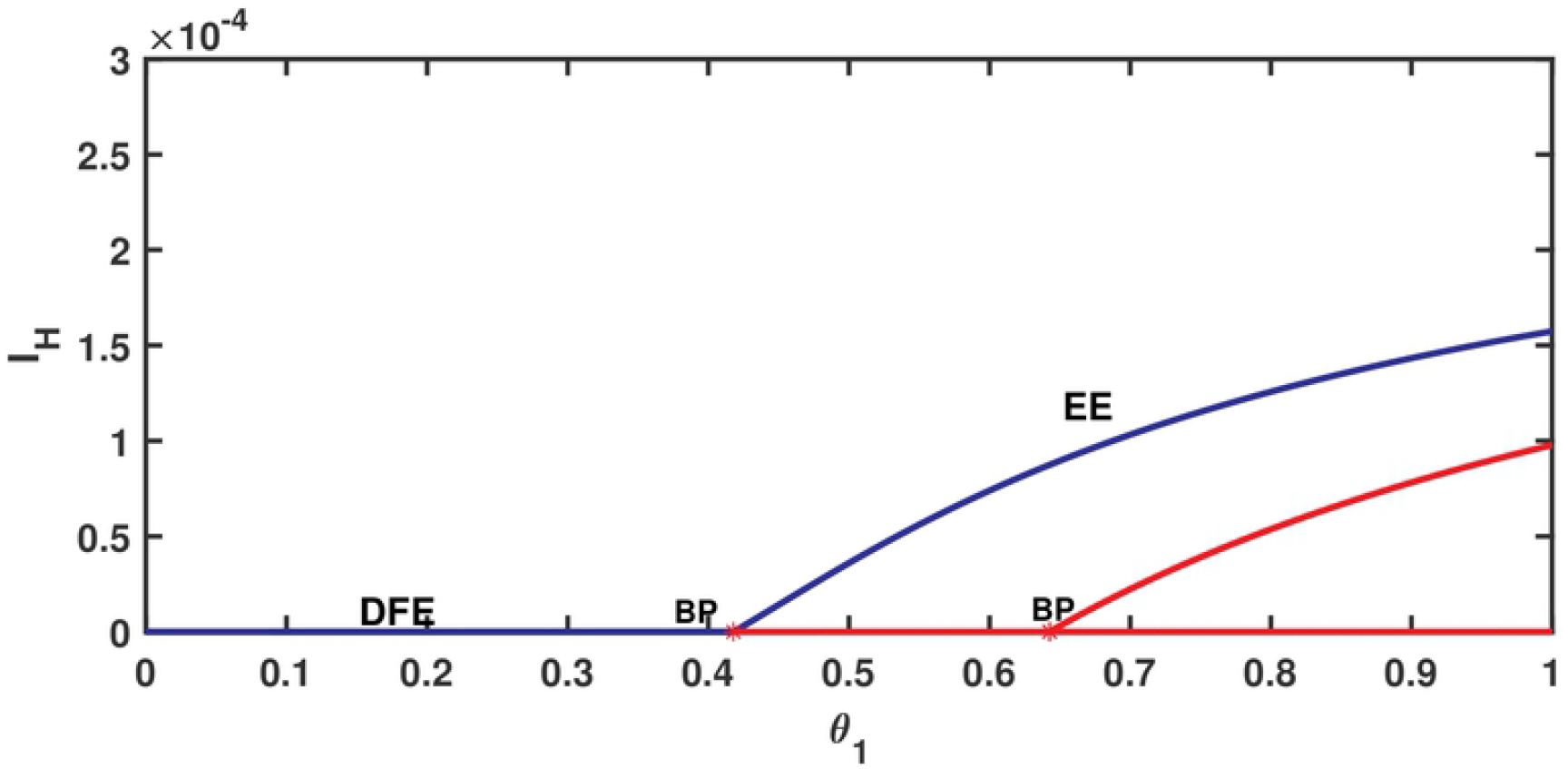
Forward bifurcation at the DFE with 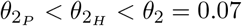 for the case ℛ_0_ = ℛ_2_ > ℛ_1_.

Next, the bifurcation result will be summarized in the following theorem and it will confirmed using Sotomayor theorem as described in [27].

#### Theorem 5

*The following holds:*

i. *If* ℛ_2_ > ℛ_1_, *then model (2) undergoes transcritical bifurcation at the DFE, given by Ƶ*^0^, *when the parameter θ*_1_ *passes through the bifurcation value* 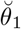.
ii. *If* ℛ_1_ > ℛ_2_, *then model (2) undergoes transcritical bifurcation at the DFE, given by Ƶ*^0^, *when the parameter θ*_1_ *passes through the bifurcation value* 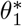*and at the AE, given by Ƶ*^(1)^, *when the parameter θ*_1_ *passes through the bifurcation value* 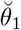.

**proof** We begin with the proof of part (i). Define the right hand side of system (2) as *Y* (*Z, t*) such that

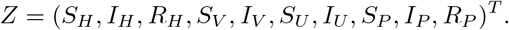

At ℛ_0_ = ℛ_2_ = 1 and ℛ_1_ *<* 1, choose *θ*_1_ as a bifurcation parameter as define in (17). The linearization matrix of system (2) around the DFE, *Ƶ*^0^ and when 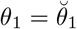 is

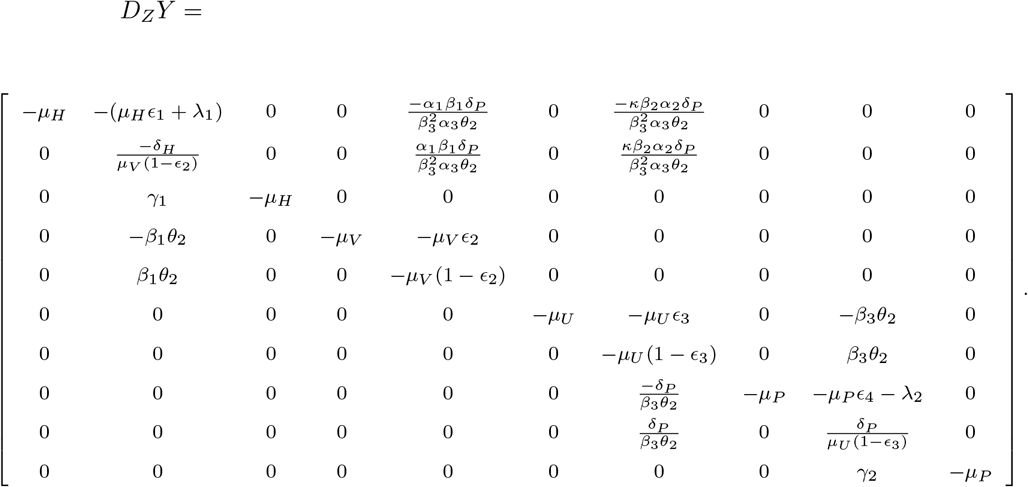

One can easily check that the above matrix has a simple zero eigenvalue.

Then, the nonzero right eigenvector *v* = (*v*_1_, *v*_2_, *v*_3_, *v*_4_, *v*_5_, *v*_6_, *v*_7_, *v*_8_, *v*_9_, *v*_10_)^*T*^ and the nonzero left eigenvector *w* = (*w*_1_, *w*_2_, *w*_3_, *w*_4_, *w*_5_, *w*_6_, *w*_7_, *w*_8_, *w*_9_, *w*_10_)^*T*^ associated with zero eigenvalue of *D*_*Z*_*Y* are

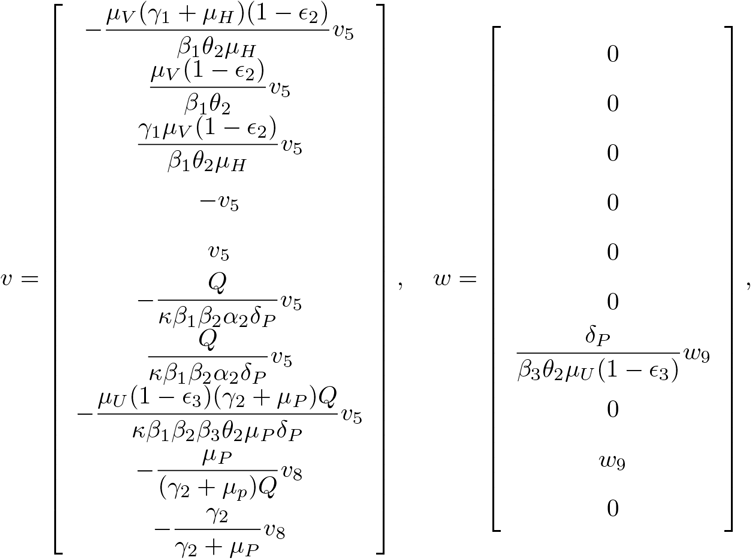

where 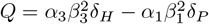. To check the first condition of Sotomayor theorem, we find the derivative of *Y* with respect to *θ*_1_ at *Ƶ*^0^ and 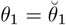, we get 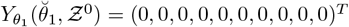. This implies

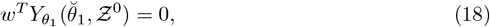

and hence first condition is satisfied. Next, we compute the Jacobian of 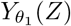 at *Ƶ*^0^ and 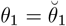 and we obtain

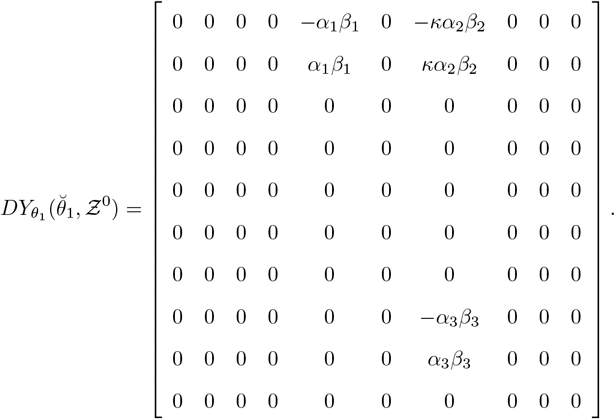

Then, we calculate

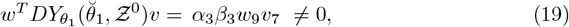

therefore, the second condition is satisfied.

Since *D*^2^ represents the matrix of partial derivatives of each components of *DY* with respect to each component of *Y* (*Z*), we get

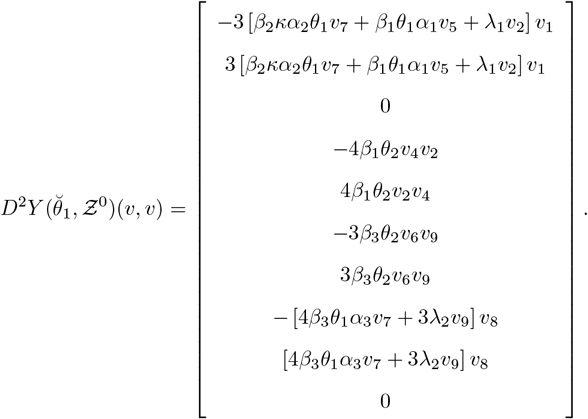

Thus, the third condition is satisfied by showing that

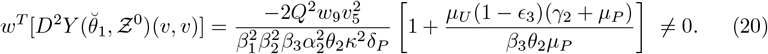

Hence, according to Sotomayor theorem, the results in (18),(19) and (20) implies that as the parameter *θ*_1_ passes through the bifurcation value 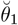 defined in (17), the system (2) experiences a transcritical bifurcation at *Ƶ*^0^ as shown on the figure above.

For the proof of part (ii), one can repeat the same procedure as we did for part (i). Choose 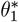 as a bifurcation parameter defined in (16). The left and right eigenvectors are

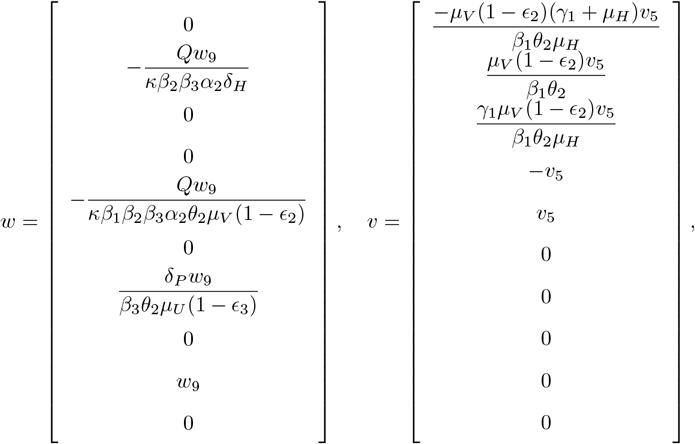

respectively. The conditions of Sotomayor Theorem hold as follows:

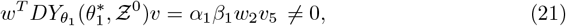

and

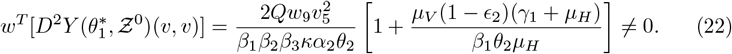

Hence, the system (2) undergoes transcritical bifurcation at the DFE when the parameter *θ*_1_ passes through the bifurcation value 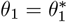. Finally, proofing the bifurcation at the AE when *θ*_1_ passes through the bifurcation value 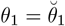can be done in a similar way.

The local stability of endemic equilibria *Ƶ*^(1)^ and *Ƶ*^(2)^ can be established from the above calculations and Theorem 4 in [15] such that *a* and *b* are given by

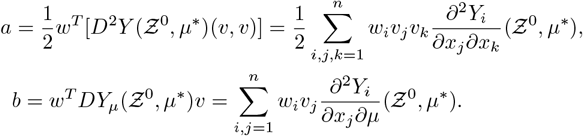

where *µ*\* is the bifurcation parameter and the result is given in the following theorem:

#### Theorem 6

*The following results hold:*

i. *Let a and b defined in (20) and (19) at* 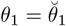 *and b ≠* 0 *(w*_9_ > 0 *and v*_5_ > 0*). Then, there exists ε*_1_ > 0 *such that the AE, given by Ƶ*^(1)^, *is locally asymptotically stable near Ƶ*^0^ *if a <* 0 *for* 0 *< θ*_1_ *< ε*_1_.
ii. *Let a and b defined in (22) and (21) at* 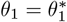 *and b* ≠ 0 *(w*_9_ *<* 0 *and v*_5_ > 0).*Then, there exists ε*_2_ > 0 *such that the EE, given by Ƶ*^(2)^, *is locally asymptotically stable near Ƶ*^0^ *if a <* 0 *for* 0 *< θ*_1_ *< ε*_2_.

The bifurcation and stability results for the case ℛ_0_ = ℛ_1_ > ℛ_2_ can be summarized as follows, taking *θ*_1_ as a bifurcation parameter and provided that the two critical values 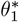 and 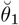 are feasible:

i. When the transmission probability from an infectious mosquito is small 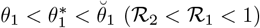, then the number of infected will decrease and eventually the disease will disappear from both areas. This implies that the DFE is stable.
ii. When 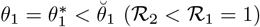, the system undergoes forward bifurcation at the DFE as *θ*_1_ pass through bifurcation value 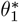.
iii. When 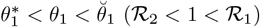, the endemic will persists only in rural area and will die out from forest area. Hence, the AE is stable, while the DFE is unstable.
iv. When 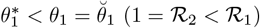, the system undergoes forward bifurcation at the AE as *θ*_1_ pass through bifurcation value 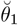.
v. When the transmission probability from an infectious mosquito is high 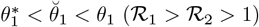, the disease will persist in both rural and forest areas and hence the EE is stable, while both the DFE and AE are unstable.

Also, we summarize the bifurcation and stability results for the case ℛ_0_ = ℛ_2_ > ℛ_1_ as follows:

I. When 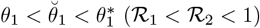, then the disease will die out from both areas and hence the DFE is stable.
II. When 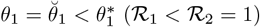, the system undergoes forward bifurcation at DFE as *θ*_1_ pass through bifurcation value 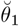.
III. When 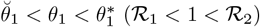, the endemic will persist in both areas and hence the EE is stable, while the DFE is unstable.
IV. When 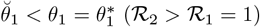, unstable AE exists as the system passes through 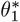.
V. When 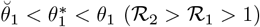, the EE remains stable and both the AE and DFE remain unstable.

#### Remark 1

*One can choose θ*_2_ *as a bifurcation parameter with the following critical values:*

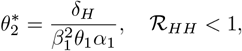

*which corresponds to* ℛ_0_ = ℛ_1_ = 1, ℛ_2_ *<* 1 *and*

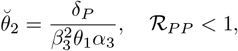

*which corresponds to* ℛ_0_ = ℛ_2_ = 1, ℛ_1_ *<* 1 *and get similar results with the feasibility of the two critical values depending on θ*_1_.

From the above discussion, we note that the stability and bifurcation results depend on both transmission probability parameters *θ*_1_ and *θ*_2_. Hence, one may also study the combined effect of *θ*_1_ and *θ*_2_. So, let us introduce a new parameter Θ defined as Θ = *θ*_1_ *θ*_2_ with two critical values 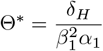 and 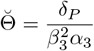. The stability region based on combined effect of *θ*_1_ and *θ*_2_ for the case ℛ_0_ = ℛ_1_ > ℛ_2_ is shown in Fig 6. From the figure, the stability and bifurcation results can be outlined as follows:

**Fig 6.**
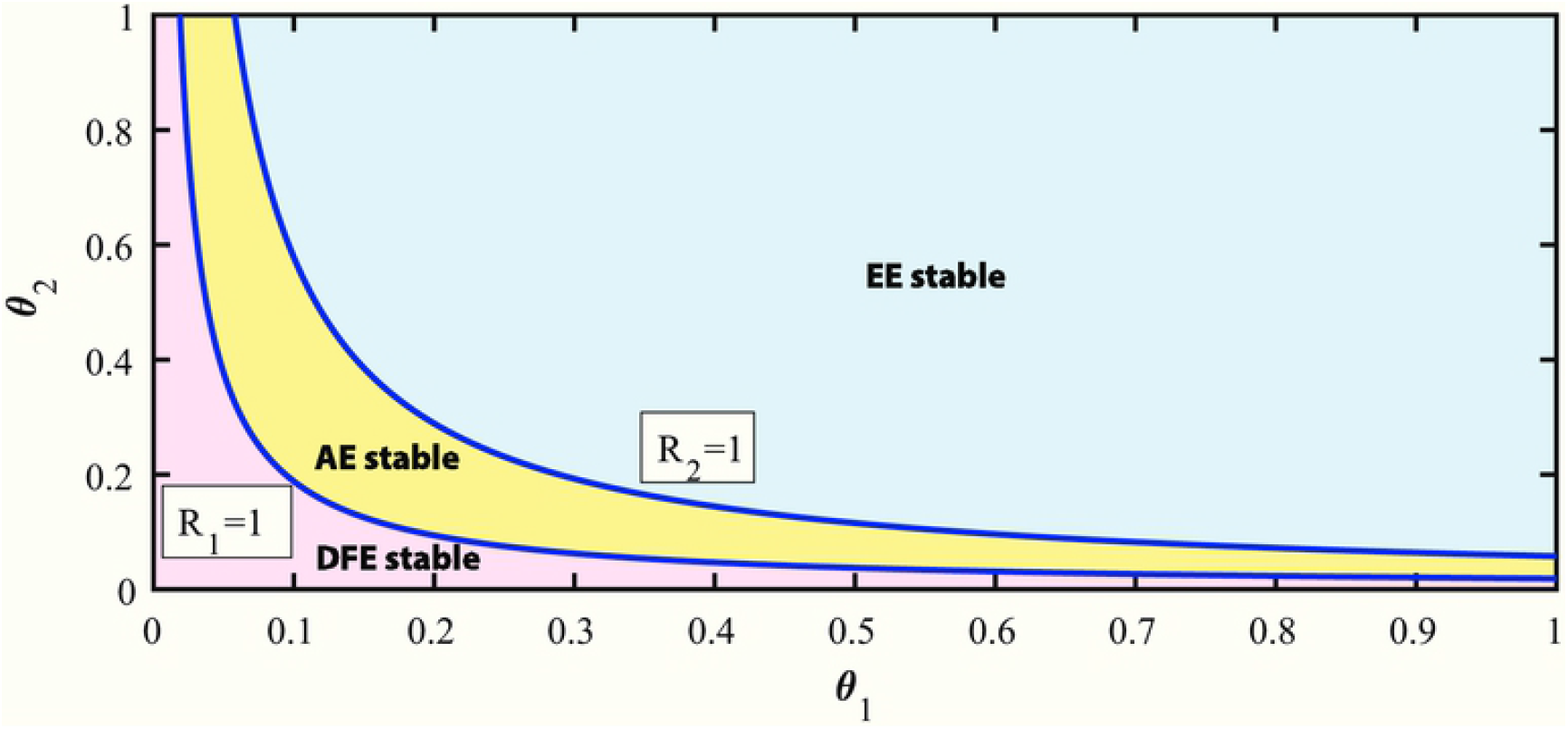
Stability region when *R*_0_ = ℛ_1_.

i. When 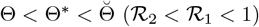, the DFE is the only stable point. Thus, the disease will die out from both rural and forest areas.
ii. The system undergoes a forward bifurcation at the DFE when Θ passes through critical value Θ*.
iii. When 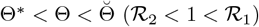, then the AE is the only stable point. Thus, the disease will persist in rural area and it will disappear from the forest area.
iv. The system undergoes forward bifurcation at the AE when Θ passes through critical value 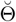.
v. When 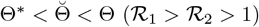, the EE is the only stable point. Hence, the disease will persist in both rural and forest areas.

The stability region of combined effect of *θ*_1_ and *θ*_2_ for the case ℛ_0_ = ℛ_2_ > ℛ_1_ is given in Fig 7. The stability and bifurcation results based on combined effect Θ for this case can be outlined as follows:

**Fig 7.**
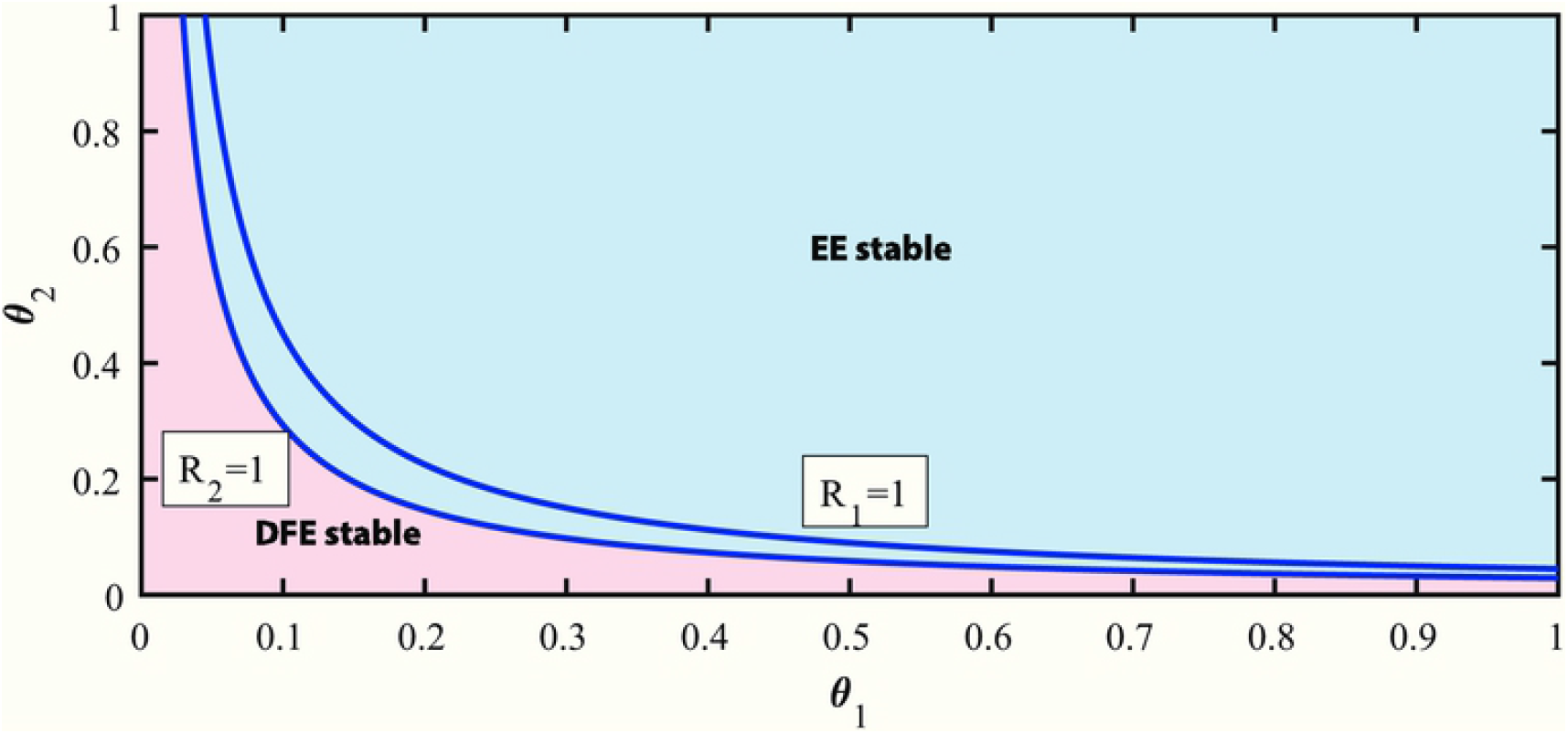
Stability region when *R*_0_ = ℛ_2_.

I. When 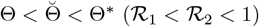, the DFE is the only stable point and the disease will die out from both areas.
II. The system undergoes forward bifurcation at the DFE when Θ passes through critical value 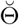.
III. When 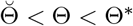 or 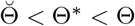, the EE is the only stable point. In other words, if ℛ_2_ > 1, the disease will persist in both areas.

### Global Stability of Axial Equilibrium *Ƶ*^(1)^

In this section, we derive the global stability of the AE. The derivation is based on finding a suitable Lyapunov function and using the LaSalle’s invariance principle. We begin by considering the Lyapunov function as follows:

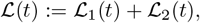

such that

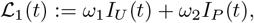

and

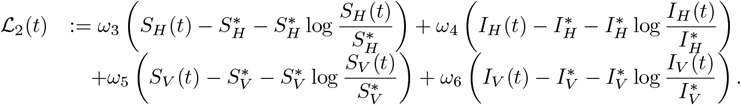

Differentiating ℒ(*t*) with respect to *t* yields:

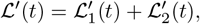

such that

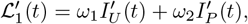

and

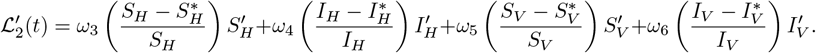

Substitution of 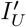 and 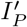 from system (2) gives

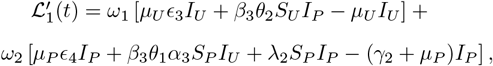

using *S*_*U*_ = 1 − *I*_*U*_ and *S*_*P*_ = 1 − *I*_*P*_ − *R*_*P*_ yields

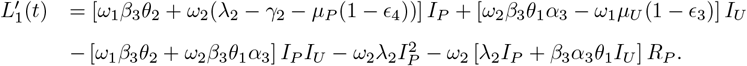

Next, choosing *ω*_2_ = 1 and 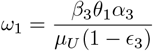 gives

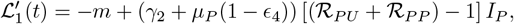

such that 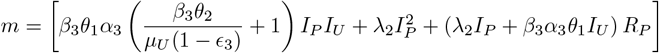.

Clearly, 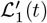 is negative definite when ℛ_2_ *<* 1.

Moreover, substituting for 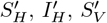, and 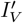 from system (2) into 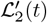 gives

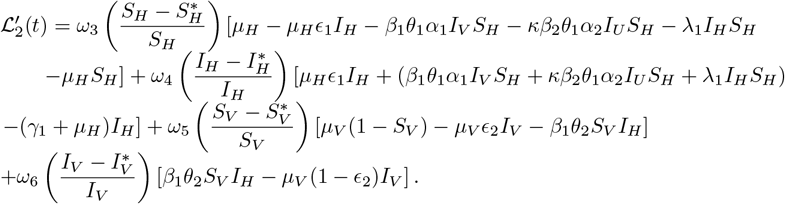

Now, let

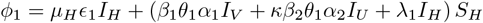

and

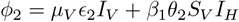

and choose

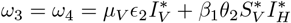

and

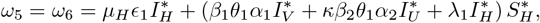

then, using the fact that 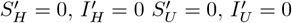 at the AE, we can write the following:

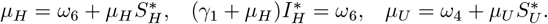

Using the above relations to rewrite 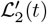, we have

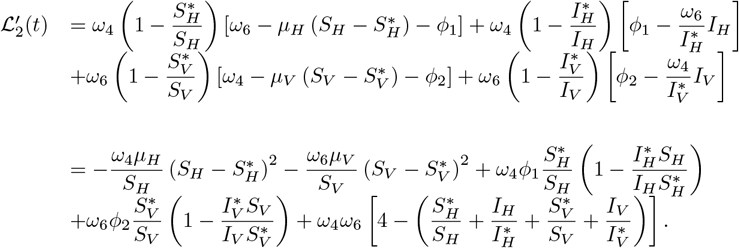

Hence, 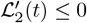 under the conditions:

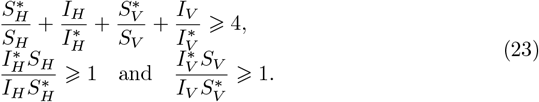

Thus, we can deduce that ℒ′(*t*) *<* 0 if ℛ_2_ *<* 1 and conditions in (23) are hold provided that ℛ_1_ > 1 which is required for existence of AE as discussed earlier. Note that ℒ′(*t*) = 0 if 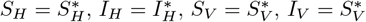 and *I*_*U*_ = *I*_*P*_ = *R*_*P*_ = 0 (*R*_*P*_ = 0, *I*_*P*_ = 0 or *R*_*P*_ = 0, *I*_*U*_ = 0). Therefore, the AE is the largest invariant set in {*X*(*t*) ∈ Ω, ℒ′(*t*) = 0}. Utilizing LaSalle’s invariance principle [28], the AE is an attractive point in Ω and hence, all solutions of system (2) converge to *Ƶ*^(1)^. The global attractivity and local stability of the AE implies that the AE is globally asymptotically stable provided that the above conditions are satisfied. This result is summarized in the following theorem:

#### Theorem 7

*The AE, given by Ƶ*^(1)^, *is globally asymptotically stable if* ℛ_2_ *<* 1, ℛ_1_ > 1 *and conditions in (23) are hold*.

### Numerical Analysis

In this section, numerical simulations are carried out to demonstrate the obtained theoretical results of ZIKV model (2) and to study the effect of some model parameters in the disease transmission dynamics. We use the base line value of the parameters as listed in Table 1 and appropriate initial conditions. The population size ratio parameters are assumed to be *α*_1_ = 2, *α*_2_ = 3, and *α*_3_ = 2.5.

### Asymptotic Behavior of the Model

The asymptotic behavior of the model is graphically verified by plotting the solution curve of human and primates populations for the same choice of initial point. Fig 8 shows that the disease is disappeared from all populations when the basic reproduction number is less than unity. Moreover, when the basic reproduction number is greater than unity, the disease either persists in rural area only as illustrated in Fig 9 or persists in both areas after exhibiting oscillatory behavior as illustrated in Fig 10.

**Fig 8.**
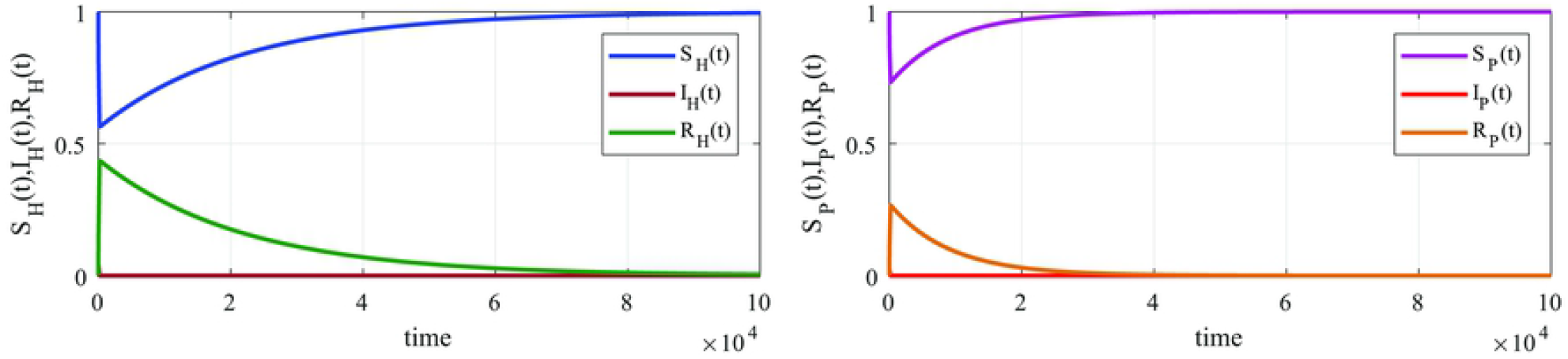
Simulation of the model (2) at ℛ_0_ = ℛ_1_ = 0.8715 and ℛ_2_ = 0.7813 for humans and primates populations, respectively.

**Fig 9.**
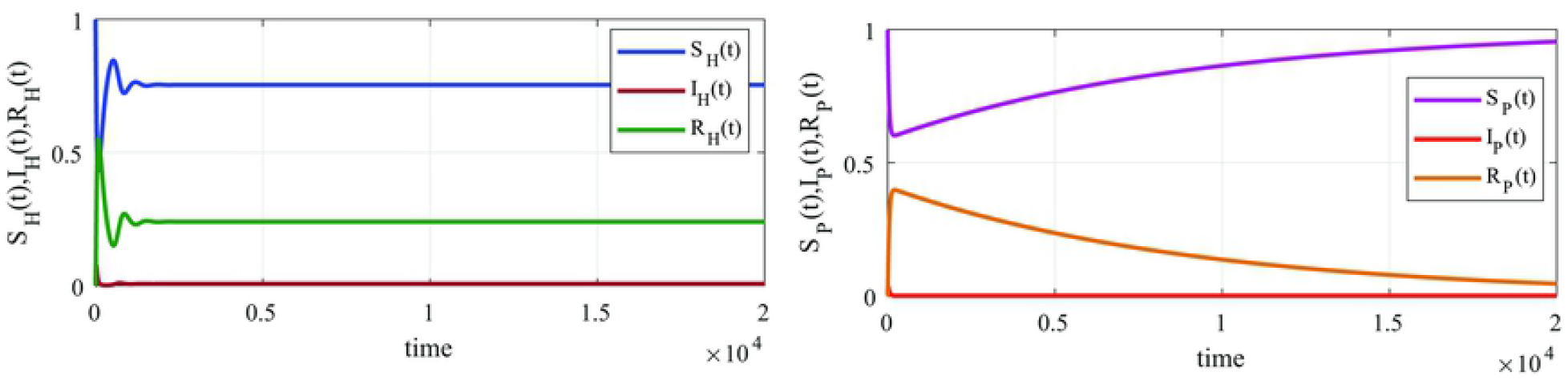
Simulation of the model (2) at ℛ_0_ = ℛ_1_ = 1.1938 and ℛ_2_ = 0.9417 for humans and primates populations, respectively.

**Fig 10.**
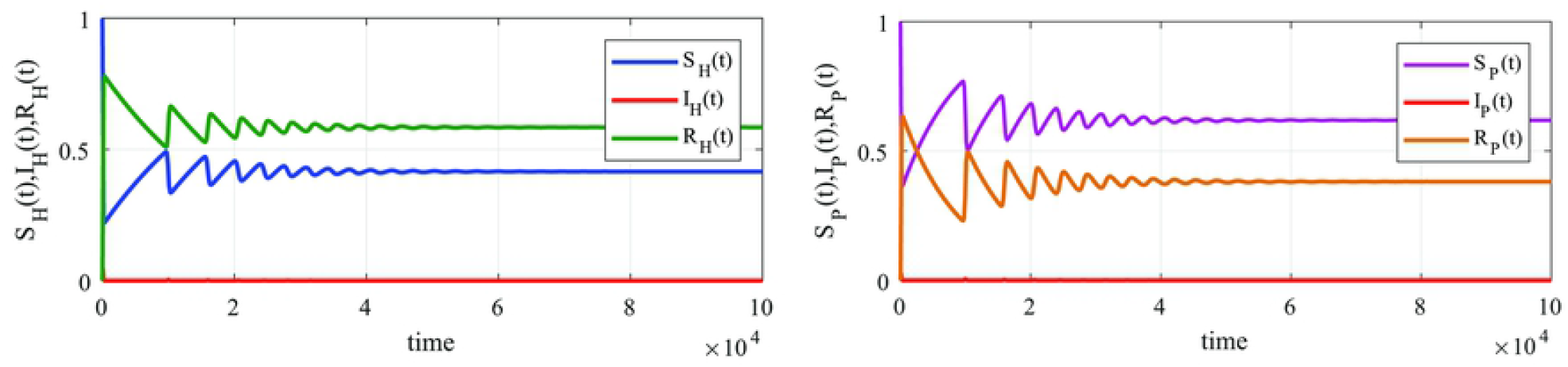
Simulation of the model (2) at ℛ_0_ = ℛ_1_ = 1.3384 and ℛ_2_ = 1.3010 for humans and primates populations, respectively.

### Effect of the Fraction of Susceptible Humans Moving from Rural to Forest Areas

Here, we investigate the effect of varying the fraction of susceptible humans moving from rural to forest areas *κ* into the number of infected human as shown in Fig 11. It can be seen that increasing the values of this rate leads to an increase in number of infected human population. On the other hand, reducing this rate has the effect of not only reducing the maximum number of infected human, but also delaying the time it takes to reach the maximum number of infection, i.e., it will have the effect of flattening the curve of infected human population. As a result, it is suggested to avoid traveling to forest area which has active virus transmission. The movement rate of humans to such areas should be targeted by control strategies in order to eliminate the ZIKV outbreak.

**Fig 11.**
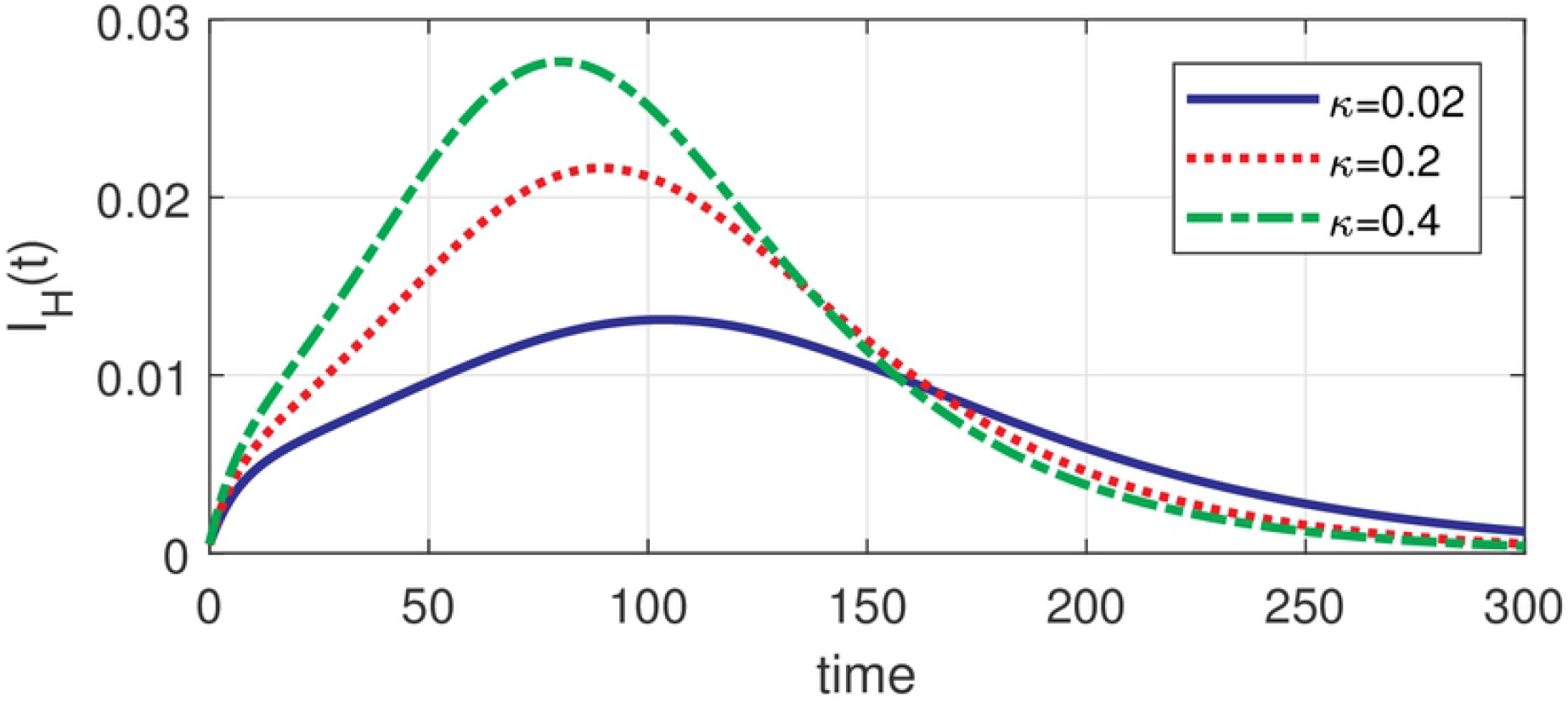
Number of infected humans for different values of *κ*.

### Effect of Varying Biting Rates

The effect of varying the values of biting rate of rural mosquitoes on humans *β*_1_, biting rate of forest mosquitoes on humans *β*_2_ and biting rate of forest mosquitoes on primates *β*_3_ on the number of infected humans are shown in Fig 12. Clearly, the biting rate of rural mosquitoes on humans *β*_1_ has the highest impact on the disease transmission among the other biting rates *β*_2_ and *β*_3_ which confirm the theoretical results of sensitivity analysis. Obviously, increasing the values of these rates will increase the number of infected human and reducing their values has the effect of flattening the curve of infected human population. The blue curve in the three graphs in Fig 12 corresponds to the case when *β*_1_ = *β*_2_ = *β*_3_ = 0.3.

**Fig 12.**
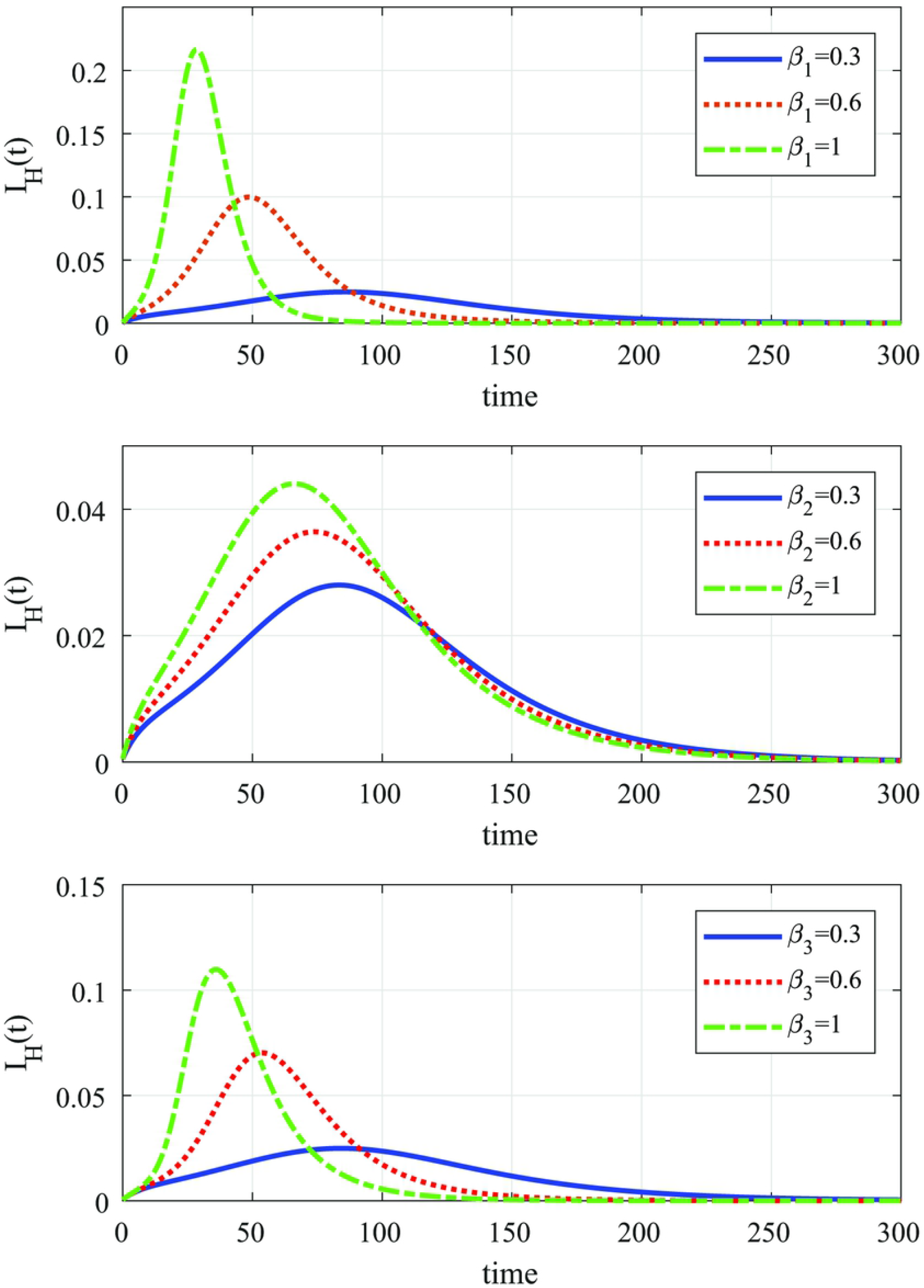
Number of infected humans for different values of *β*_1_ (Top), *β*_2_ (Middle) and *β*_3_ (Bottom).

Now, comparing the effect of *β*_2_ and *β*_3_, one could easily see from Fig 12 that the indirect impact of *β*_3_ on infected humans is higher than the direct impact of *β*_2_, which is expected since the infectious primates is considered to be the only source of infection for forest mosquitoes and hence the higher the biting rate on primates the more they become infectious to human even with fixed biting rate on human. On the other hand, fixing the biting rate on primates at low values, taken here to be *β*_3_ = 0.3, the forest mosquitoes will have less impact on infected human even with increasing biting rate on human. This shows that interaction between forest mosquitoes and primates, represented here through *β*_3_, has a significant impact on the spread of ZIKV among human population when humans travel to forest areas from some reasons. Therefore, as mention previously, control strategies should impose restrictions on travel to affected forest areas of ZIKV in order to control the spread of the disease among the human population.

### Combined Effect of Transmission Probabilities

Here, we present the effect of transmission probabilities of infection from and to mosquitoes, *θ*_1_ and *θ*_2_, respectively. Their effect on human and primate populations is illustrated in Figures 13-14. We vary both *θ*_1_ and *θ*_2_ and fix the values of all other parameters for the two cases (a) ℛ_0_ = ℛ_1_ > ℛ_2_ and (b) ℛ_0_ = ℛ_2_ > ℛ_1_. The values of *θ*_1_ and *θ*_2_ are chosen based on the critical values of their product Θ = *θ*_1_*θ*_2_. Fig 13 illustrates their effect for the case (a) with critical values Θ* = 0.0188 and 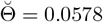,. It can be seen that when the combined transmission probability is small,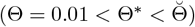, both human and primate populations converge to the DFE and hence the disease dies out from both rural areas and forest areas. When 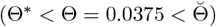, the human population reaches the endemic steady state after exhibiting oscillations in the susceptible human population, while the primate population goes to the disease free steady state. This agrees with the obtained stability and bifurcation results, i.e., the system converges to the stable AE for this case. Hence, the disease will persist in rural area and will disappear from forest area. For higher combined transmission probability 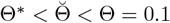, both populations reach the endemic steady state after exhibiting oscillation in the susceptible populations. Hence, the disease will persist in both areas. Fig 14 is devoted for case (b) with the critical values of Θ are 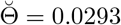 and Θ* = 0.0449. When 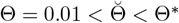, the system reaches the stable DFE. However, if the combined transmission probability is 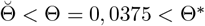 or 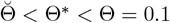, the system reaches the stable EE after exhibiting oscillation behavior as shown in Fig 14. Reaching a stable AE is not feasible in this case which is consistent with bifurcation results since the system undergoes forward bifurcation only as Θ passes through critical value 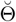.

**Fig 13.**
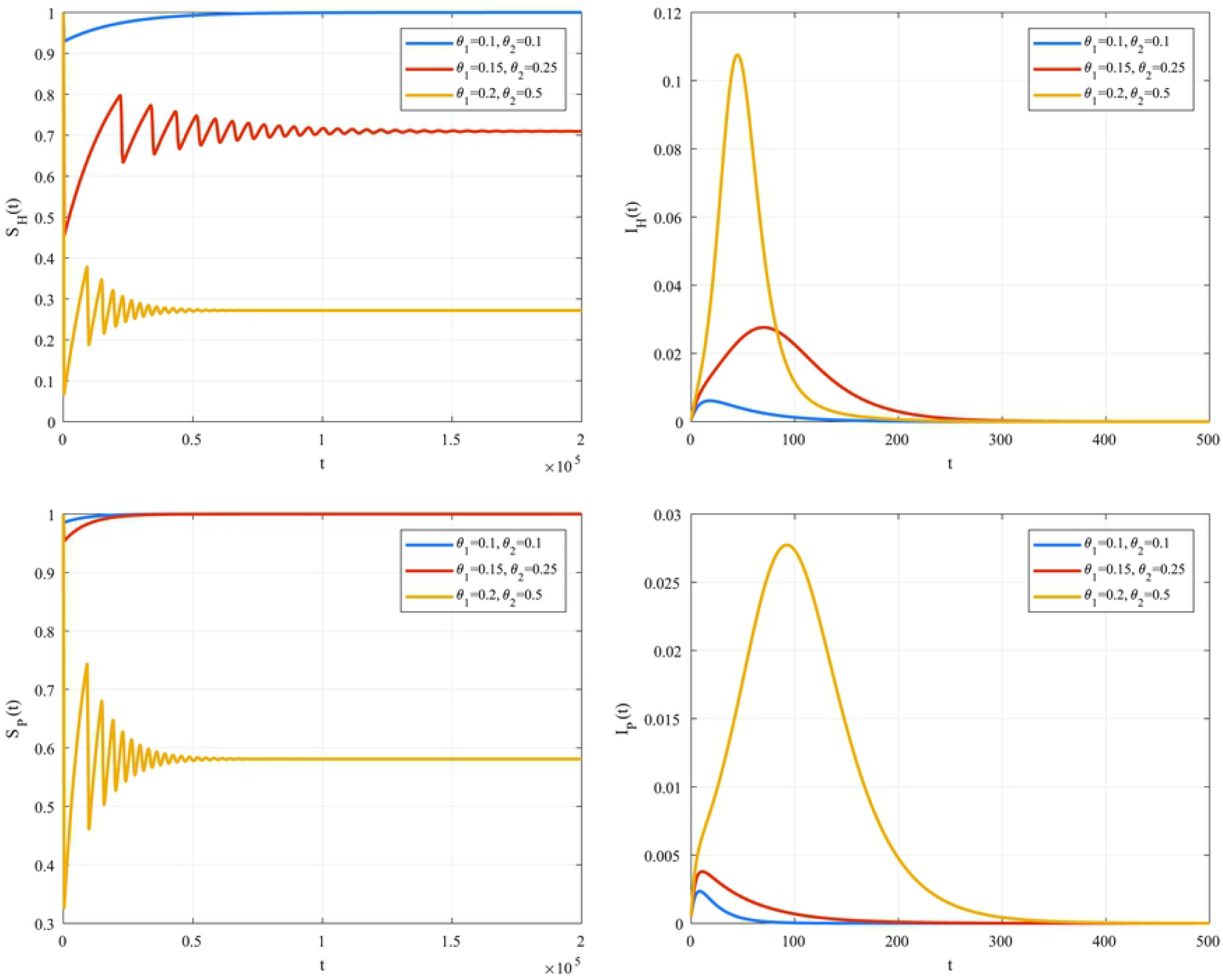
Combined effect of *θ*_1_ and *θ*_2_ on human and primates populations for the case (a) ℛ_0_ = ℛ_1_ > ℛ_2_.

**Fig 14.**
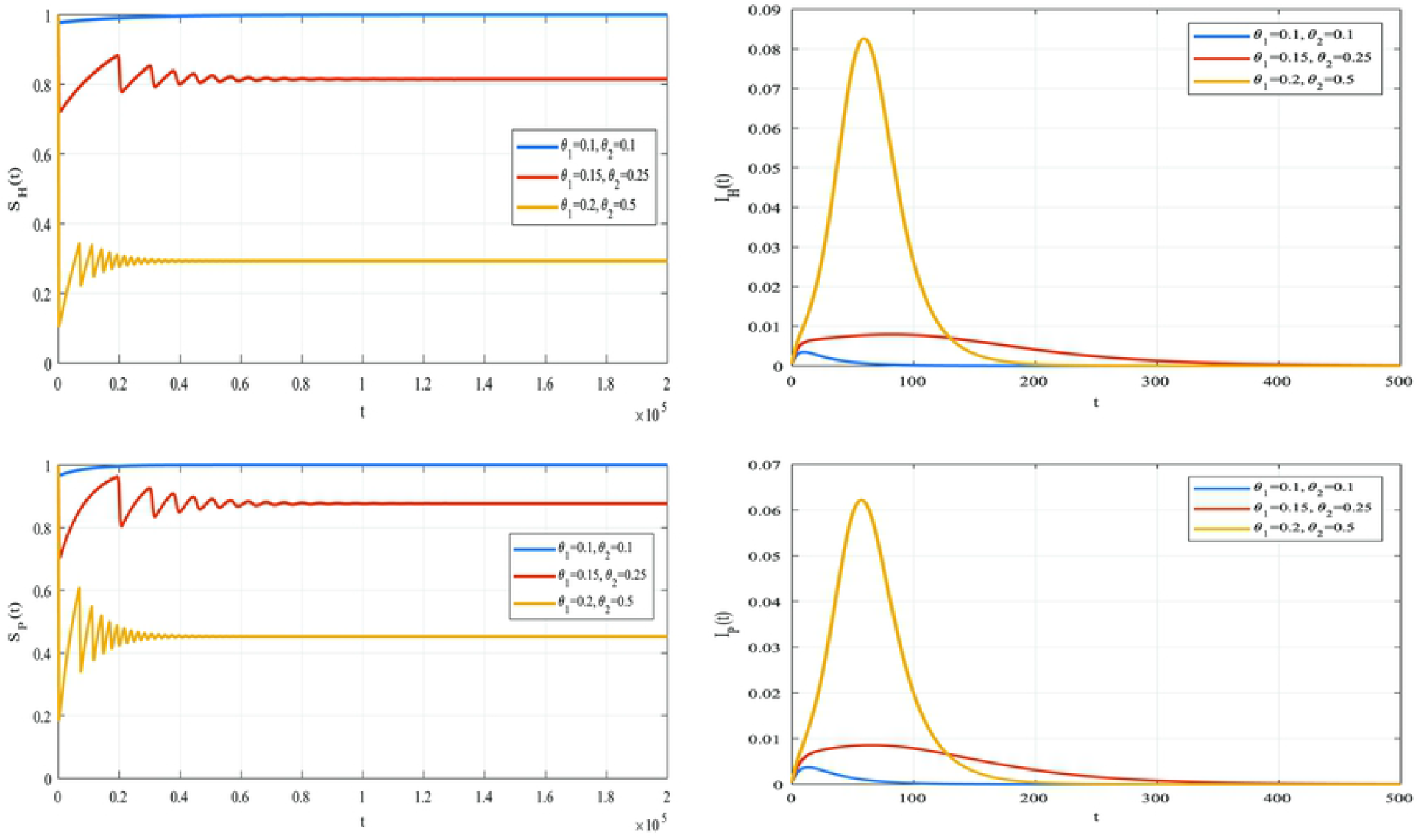
Combined effect of *θ*_1_ and *θ*_2_ on human and primate populations for case (b) ℛ_0_ = ℛ_2_ > ℛ_1_.

## Conclusion

A mathematical model of ZIKV illness including direct and vertical transmission of human, mosquito vectors and monkey primates has been proposed. Human movement from rural area to forest area has been also considered. The model was studied and fully analyzed to investigate the impact of primates and human mobility from rural area to forest area in the spread of ZIKV. The boundedness of the invariant region and the positivity of the solution were discussed. The basic reproduction number ℛ_0_ was computed and expressed in terms of two terms ℛ_1_ and ℛ_2_, where ℛ_1_ represents the transmission due to the interaction within human population ℛ_*HH*_ and the interaction between humans and vectors in rural areas ℛ_*HV*_, whereas ℛ_2_ represents the transmission due to the interaction within primate population ℛ_*PP*_ and the interaction between primates and vectors in forest areas ℛ_*PU*_. The disease threshold occurs either whenever ℛ_*HH*_ + ℛ_*HV*_ = 1 if ℛ_0_ = ℛ_1_ = 1 or whenever ℛ_*PP*_ + ℛ_*PU*_ = 1 if ℛ_0_ = ℛ_2_ = 1. The proposed model were found to have three equilibria: a disease free equilibrium (DFE), which always exists, an axial equilibrium (AE) exists when ℛ_1_ > 1 and it describes an endemic in populations living in rural area only and a full endemic equilibrium (EE) describing an endemic in both forest and rural areas, which exists when ℛ_2_ > 1. Then, sensitivity analysis of ℛ_0_ was carried out, which revealed that ℛ_0_ is sensitive to practically all model parameters, either positively or negatively. However, the most positive influential parameters are the biting rate of rural mosquitoes on humans and the biting rate of rural mosquitoes on primates, while the recovery rate of humans has the most negative impact. The sensitivity analysis results also showed that the transmission probabilities from and to mosquitoes, *θ*_1_ and *θ*_2_, respectively, are as important as the ratios of population size between vector and human populations, *α*_1_, and between vector and primates populations, *α*_3_, in the disease transmission dynamics. The effect of the transmission probabilities has been further investigated in the bifurcation analysis and through numerical simulation. The bifurcation analysis was investigated analytically and graphically for the case when ℛ_0_ = ℛ_1_ > ℛ_2_ and the case when ℛ_0_ = ℛ_2_ > ℛ_1_, taking one of the transmission probabilities as a bifurcation parameter. The feasibility of forward bifurcation were found to depend on the values of both transmission probabilities *θ*_1_ and *θ*_2_. It was observed that if ℛ_0_ = ℛ_1_ > ℛ_2_, the proposed model may have a double forward bifurcation at DFE and AE, respectively, while if ℛ_0_ = ℛ_2_ > ℛ_1_, it may only experience a forward bifurcation at DFE. It was shown that the DFE is globally asymptotically stable whenever ℛ_0_ is less than unity, the AE is globally asymptotically stable whenever ℛ_1_ > 1 > ℛ_2_ under certain conditions and the EE is locally asymptotically stable whenever ℛ_2_ > 1 since the direction of the bifurcation was found to be forward. The transmission probabilities were found to play a critical role in the obtained stability results. Finally, Numerical simulations were carried out to demonstrate the obtained theoretical results and to study the effect of some model parameters in the disease transmission dynamics. In particular, it was illustrated that the model converges asymptotically to the DFE whenever ℛ_0_ *<* 1, to the AE whenever ℛ_1_ > 1 > ℛ_2_ and to the EE after exhibiting oscillatory behavior whenever ℛ_2_ > 1. It was also illustrated that the interaction between forest mosquitoes and primates has a significant impact on the ZIKV transmission dynamics among human population and that reducing the fraction of susceptible moving to forest areas has the effect of flattening the curve of infected human population. The critical impact of the combined transmission probabilities from and to mosquitoes in the disease dynamics has been also illustrated numerically. In particular, it was shown that for high transmission probabilities, the disease presists in all populations after exhibiting oscillatory behavior.

## Data Availability

All relevant data are within the manuscript and its Supporting Information files.

## Notes

### Competing Interest Statement

The authors have declared no competing interest.

### Funding Statement

The author(s) received no specific funding for this work.

